# Increased airborne transmission of COVID-19 with new variants, Implications for health policies

**DOI:** 10.1101/2022.01.13.22269234

**Authors:** Bertrand.R. Rowe, André Canosa, Amina Meslem, Frantz Rowe

## Abstract

New COVID-19 variants, either of higher viral load such as delta or higher contagiousness like omicron, can lead to higher airborne transmission than historical strains. This paper highlights their implications for health policies, based on a clear analytical understanding and modeling of the airborne contamination paths, of the dose following exposure, and the importance of the counting unit for pathogens, itself linked to the dose-response law. Using the counting unit of Wells, i.e. the quantum of contagium, we develop the conservation equation of quanta which allows deriving the value of the quantum concentration at steady state for a well-mixed room. The link with the monitoring concentration of carbon dioxide is made and used for a risk analysis of a variety of situations for which we collected CO_2_ time-series observations. The main conclusions of these observations are that 1) the present norms of ventilation, are both insufficient and not respected, especially in a variety of public premises, leading to high risk of contamination and that 2) air can often be considered well-mixed. Finally, we insist that public health policy in the field of airborne transmission should be based on a multi parameter analysis such as the time of exposure, the quantum production rate, mask wearing and the infector proportion in the population in order to evaluate the risk, considering the whole complexity of dose evaluation. Recognizing airborne transmission requires thinking in terms of time of exposure rather than in terms of proximal distance.

**Highlights:**

- Relative airborne risk assessment following variant viral load and contagiousness
- Indoor analytical risk assessment including absence of ventilation
- Adequacy of the present norms of ventilation to Covid-19 pandemic
- Observation of non-compliance to standards concerning CO_2_ Indoor Air Quality

## I Introduction

Since its emergence at the end of 2019 a variety of public and health measures and recommendations have been decided in several countries to contain COVID-19 spreading. Recommendations pertain more to personal hygiene as, for example, washing hands, coughing in his elbow, and keeping a social distancing with other individuals. However, collective measures have been often more coercive. They include, amongst others, lockdown, closing of specific activities such as restaurant services, quarantine, sanitary pass and last but not least human surveillance data tracking. These mitigation measures have often had profound side effects, sometimes deleterious, on the economy and population mental health [1].

Developing a rational basis for prevention is necessary to avoid irrational measures such as forbidding outdoor activity in under-crowded area or organizing a kind of carousel circulation in commercial centers. This requires identification of causal mechanisms, i.e. risk factors, explaining the spread of the disease. A rational public health policy requires careful evaluation of the pharmaceutical and non-pharmaceutical interventions. This should be the key role of epidemiology [2].

As described in a large number of publications, there are three routes of transmission of respiratory diseases. The first can be considered as a person-to-person transmission, occurring via direct close contact, when microdroplets of physiological fluids emitted by an infected person are projected directly on the mucosa (lips, nose, eyes) of another person in a kind of ballistic way. The second one is linked to self-touch of the face mucosa by hands contaminated by surfaces ((fomites) or projections. The third route, known as “aerosol” or “airborne”, is due to the creation of a persistent aerosol of microdroplets in a range of size which prevents their rapid sedimentation on the floor. This aerosol emitted by an infected person can be re-breathed leading to further contamination. Mainly due to historical reasons [3] it was outright denied by most of health authorities including WHO, or governmental agencies such as the CDC in the US (Center of Disease Control) or the HAS (Haute Autorité de Santé) in France. Then mitigation measures were decided considering the first two ways of transmission: social distancing, washing hands etc. Unfortunately for public health, the consideration of airborne transmission should have led to a variety of other decisions, especially in the field of indoor air quality (hereafter IAQ).

Ironically, knowledge was available for suspecting the importance of airborne transmission in the COVID-19 pandemic. As soon as the first half of the last century, Wells and his co-workers have led numerous experiments and developed concepts still largely in use nowadays in the field of respiratory diseases. Wells has exposed his visionary ideas and summarized his work in a book of 1955 that any epidemiologist should have read [4]. With his coworker Riley he developed the famous Wells-Riley model [5] which has been the basis of a lot of avatars and developments, especially in the last two decades [6,7].

The non-consideration of airborne transmission has led L. Morawska, a leading scientist in the field, to raise an alarm on its importance [8], followed by a call co-signed by more than two hundred researchers in the mainstream press [9]. Nowadays the very importance of airborne transmission of the COVID-19 disease is largely recognized and the reader is referred to the review in Science (and references therein) of Wang et al [10], leading to the conclusion that airborne transmission is the major spreading route. Complementary details can be found in [11-16].

Viruses mutate constantly, leading to new variants, eventually more infectious than the previous strains, modifying the epidemiology of the disease. Variant classification is beyond the scope of the present paper and rather complicated since there is not a single nomenclature. Their scientific name refers to their lineage (a lineage is a group of closely related viruses with a common ancestor) and to mutations resulting from changes in the genetic code leading eventually to new variants [17]. An expert group of WHO has recommended using letters of Greek alphabet to name variants in non-specialized audience [18]. Recently it was shown that the δ variant (B.1.617.2), which appeared first in India in October 2020, leads to a much higher viral load (hereafter VL) in respiratory fluids than initial strains, referred hereafter as IS [19-21]. According to recent observations, the new omicron variant, spreading very fast in a number of countries, has a smaller VL than the δ one but is nevertheless more contagious for microbiological reasons.

We rationalize below why new variants lead to a much higher airborne transmission, essentially for the case of homogeneous transmission in indoor environment, following the Wells-Riley approach. The relative risk for different variants (following VL and microbiological characteristics), and various situations, is calculated. We have also performed measurements of CO_2_ concentrations in a variety of environments, demonstrating that in the real-life ventilation is seriously insufficient and that the homogeneous hypothesis is most often verified. We finish by emphasizing the implications for health policy of the increased airborne transmission, which is certainly the main transmission way for new variants. Following other authors [22] we insist on the importance of **the time of exposure** although unfortunately most of the public policy is based on the **distance of exposure**, probably due to the initial denial of airborne transmission.

## II basic notions and models in airborne transmission

### II-1. Infectious particles and VL

Particles emitted by a human refer either to spherical microdroplets or to more or less hydrated “dry nuclei”, resulting from water evaporation of the respiratory fluids, which, beside water, contains minor components like mucus, proteins and viruses [23]. VL is a key parameter of particle infective power and depends on the mean number of viruses per unit volume of respiratory fluids, which lead to a mean number per particle. This latter is statistical, i.e. it implies a large distribution of particles with various viral contents. A mean VL per particle lower than unity implies that some microparticles will contain a virus and others will not. Moreover, evaporation of exhaled microdroplets can result in particles of lower size without virus loss. Since the smallest particles are very abundant, they can be very efficient in airborne transmission.

These particles can be characterized by their size and composition, including VL which depends on the viral strain. Their size depends mainly on their origin from the respiratory tract and of their evolution in the ambient air, including evaporation. The largest droplets, behaving in a ballistic way, are most often emitted by talking, sneezing, or coughing. The smallest ones come from various parts of the respiratory tract, including the lungs. They have a large distribution of sizes, and many are below 10 μm, especially after evaporation of some of the largest ones. In a kind of reversible way, the smallest ones (< 5 μm) can penetrate deep in the lung when re-breathed and are known as respirable aerosols [10,24].

One of the most sophisticated apparatuses used for the size characterization of these aerosols is the specific wind tunnel developed by L. Morawska and her coworkers at the Queensland University of Technology, at Brisbane, in Australia. It uses a variety of sizing techniques [25,26]. They found four main modes in the distribution of particle size, centered around 0.8, 1.8, 3.5, and 5.5 μm respectively.

### II-2. Concepts of dose and quantum

As discussed in Rowe *et al*. [6] and others [27,28] the notions of level and dose of exposure are easily defined for chemical or physical hazards (such as toxic gases or asbestos): the level of exposure is then the concentration of toxic and the dose the quantity inhaled, ingested etc. These definitions are much more difficult for biological pathogen agents that are not easy to measure and have the possibility to replicate in the target host [28,29]. Concerning aerosols and as stated by Haas *et al*. [28] “precise information on the concentration of pathogens in aerosols has a lot of uncertainty associated with it”. Moreover, and for any kind of disease (i.e. respiratory, digestive etc.), the effect of the dose could depend on the way of transmission: inoculation, ingestion, airborne etc. Having defined a dose, the work of epidemiology is to assess quantitatively the risk for a given dose: by nature, such an assessment is statistical; it results most often in a law linking the probability of infection to the dose.

For airborne transmission of respiratory diseases, the definition of a dose is far from being straightforward since measuring pathogen concentrations in the air is extremely difficult [28]. Therefore, **Wells** [4] **defined the quantum of contagium** as a hypothetical quantity that has been inhaled per susceptible individuals (men or animals) when 63.2% (correspondingly to a Poisson dose-response law, see sub-section II-4) of these individuals display symptoms of infection. Quantum is used throughout the present paper and contrary to what has been sometimes claimed [30], it has no dimension **but is a counting unit** (like moles compared to molecules). It considers a variety of mechanisms: inhalation of airborne particles, pathogen inhibition by host defenses (see supplementary materials1, hereafter SM1-7) or other losses, before any replication will start in an infected cell. Therefore, it corresponds statistically to a number of pathogens higher than one.

However, these statistical concepts do not mean that very few pathogens are never enough to start infection, as assumed sometimes. Indeed, the so-called “single hit” models make statistical risk assessment considering a very small probability, although non-zero, of infection by a single pathogen [28,31-34]. Further, and as stated by Haas *et al*. [28], the term of Minimum Infective Dose is very misleading since “Minimum” suggests some threshold effect for the infection. They emphasize that it corresponds in fact to the average dose administered and most frequently relates to the value required to cause half of the subjects to experience a response; they suggest that “median infectious dose” should be more appropriate, and they show that it is not possible to infer the probability of infection by a single pathogen from the magnitude of the median infectious dose.

### II-3. Link between the quantum production rate and infectious aerosols

Evaluation of quantum concentration in air requires knowing the production rate of quanta by an infector, defined per unit time (unit: h^-1^ for example). It can be deduced from epidemiological observations [35] but also linked to the distributions of microdroplets emitted by humans, together with the knowledge of VL in respiratory fluids and of the efficiency of the viral strain.

Following Buonanno *et al*. [36] the production rate of quanta *q* can be written as:

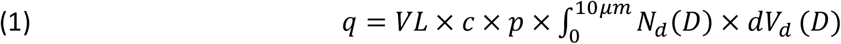

where *VL* refers to unit volume viral load of respiratory fluid, *c* is a proportionality factor between the exhaled viral content (copies/unit time) and quanta, *p* is the pulmonary exhaled volume rate (volume/unit time), *N*_*d*_(*D*) the size distribution of droplet concentration (diameter *D*) of volume *V*_*d*_. The factor *c* depends on the microbiological characteristics of the variant and can explain a higher value of *q* (and hence a higher contagiousness) even with a lower *VL*.

Equation (1) implies that **the production rate of quanta can be considered as proportional to VL in the respiratory fluids and to a factor (c) which depends on the virus microbiological characteristics**. Equation (1) assumes a single mean value of VL. This is a reasonable assumption since the quantum production rate is a statistical mean quantity that does not consider the diversity of particle emission processes, although VL depends probably on the particle origin from the respiratory tract. Note also that the integral in (1) is just the volume fraction of emitted microdroplets.

### II-4. Dose calculation and infection probability

In absence of masks the dose of inhaled quanta can be expressed as the integral over time of exposure of the product of quantum concentration *n*_*q*_ (quanta per unit volume) by the pulmonary volume inhalation rate *p* (volume per unit time):

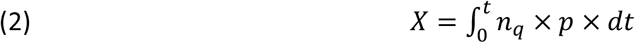

Note that this definition does not require a homogeneous distribution of quanta in space. Only 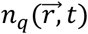 at mouth and nostrils location has to be considered. Also due to the extremely low concentration of quanta in air, 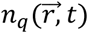 is not really continuous but can be treated as such due to the statistical aspect of the problem (as discussed previously for the VL of microdroplets).

This dose *X* has no dimension but is dependent of the choice of the counting unit with its dose-response (probability) function, which, for quanta, is the Poisson law:

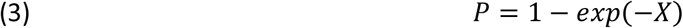

For *X* ≪ 1, this probability of transmission is then just *X*.

There are several other dose-response functions and dose definitions that can be used [27,28,37]. In any cases, the probability of infection must be a monotonically increasing function of the dose, starting from zero at dose zero and increasing toward an asymptote *P* = 1 at large dose.

### II-5. models of transmission

Whatever the chosen counting unit for the pathogens (viruses, quanta, particles), dose evaluation requires to determine spatio-temporal evolution of their concentrations. For quanta it is possible to distinguish between homogeneous models for which:

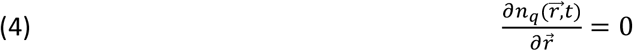

and inhomogeneous ones which consider the possible gradients of *n*_*q*_ in space:

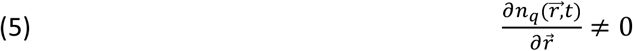

In both cases the determination of *n*_*q*_ evolution uses conservation equations, described in SM2, together with the well-mixed room hypothesis employed in homogeneous models.

The temporal evolution of quantum concentration in the homogeneous case reads (see SM2):

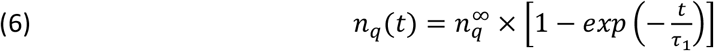

with:

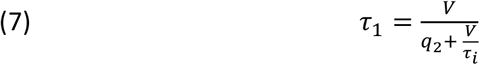

*V* being the room volume, *q*_2_ the room ventilation rate and *τ*_*i*_ the virus lifetime.

The concentration of quanta for a number of *I* infectors, at stationary state *i*.*e*. for *t* ∼ a few *τ*_1_is:

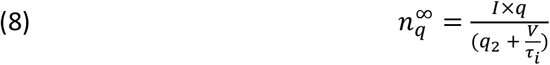

which, if the virus lifetime is long enough, reduces to:

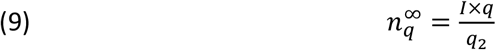

Note that if there is some air treatment (filtration or sterilization or both) for the volume *V*, it can be considered as an increase in the flow rate of fresh air and therefore results in an increase of *q*_2_ value. Indeed, it is also possible to introduce the virus lifetime as an increase in the ventilation flow rate through equations (7) and (8). The virus lifetime *τ*_*i*_ depends on a variety of phenomena including UV irradiation.

In a situation where the stationary state has already been reached in a homogeneous volume at the beginning of exposure then, following equation (9 and 2), the inhaled dose is:

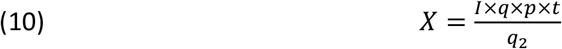

which yields for the probability of transmission:

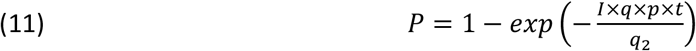

Together with the quantum definition, these equations are the basis of the Wells-Riley model [5]. Note that conservation equation (see SM2) allows to consider any unsteady cases, including the case of very poorly ventilated rooms which is equivalent to *q*_2_ ≪ *V*/*t, t* being the time of exposure. Then, assuming a zero quantum concentration at *t* = 0 (case of a tutorial room at the beginning of a lecture after a weekend for example) the dose of exposure now reads:

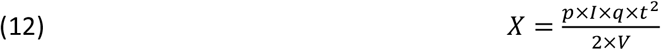

which is valid at 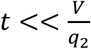 and can be used with the Poisson probability law.

In many circumstances homogeneous models are completely relevant to indoor situations, as shown by measurement of CO_2_ used as an indicator, or by considering turbulent indoor flow with typical velocities around 0.1-0.2 m/s induced by natural or mechanical ventilation or by air movement due to plumes from occupants or any hot surface. However, there are undoubtedly conditions where substantial gradients of pathogens (quantum) prevail leading to a risk which is dependent on the indoor position of infectors and susceptible persons. Two situations can be depicted for inhomogeneous transmission: the case of indoor viral transport on rather large distances, i.e., which are close to the space typical length [22] and the event of close contact between an infector and a susceptible person [37]. The concepts described above for homogeneous models are still valid but now the determination of 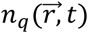 requires solving transport equations as described in SM2. Note that It is now largely admitted that the transmission of COVID-19 disease by close contact is most often an airborne one, referred in the literature as “short-range airborne transmission”. In their paper, Cortellessa *et al*. have also considered large microdroplets which have ballistic trajectories and shown that they prevail only at very short distance (< 60 cm), with a contribution to the dose being completely negligible further. This demonstrates the airborne character of most airborne contamination in close contact, excepted intimate. Other implications of this work are found in SM3.

## III Relative risk assessment following variant VL and contagiousness

### III-1. General formulation

As developed previously, airborne models of infection usually introduce a dose of exposure *X* to an infective agent, which is assumed proportional to VL in the respiratory fluids. Then the probability of infection follows a dose-response function.

**All other parameters being equal** (time of exposure, flow rate of fresh air etc..), it is then possible to assess a relative risk between two variants (in a way similar to Rowe *et al*. [6] for the relative outdoor versus indoor risk). For sake of simplicity, we concentrate the following discussion on the initial strain and the δ variant with different VL, *VL*_*IS*_and *VL*_*δ*_ respectively.

Let *R* be the ratio of the doses of exposure between *IS* and δ in case of identical situations, from section II (Eq. 1 and 2), *R* can be reduced to the ratio of VLs and of the proportionality factors c:

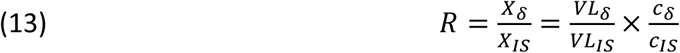

It is then easy to demonstrate that relative probabilities of being infected between respectively δ and *IS* variants follow the next equation:

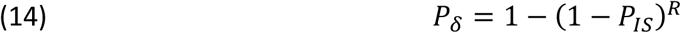

which for *P*_*IS*_ ≪ 1 reduces to *P*_*δ*_ = *R* × *P*_*IS*_.

It results that, from the recognized fact that *VL*_*δ*_ ≫ *VL*_*IS*_, the airborne contamination by the δ variant is much more efficient than with initial strains for comparable situations, as shown in Figure 1 for *R* = 10 and 100 respectively. Note that the same conclusion could apply with the omicron variant (the subscript *δ* should be replaced by *o*) but then the *c*_*o*_ factor would also explain the higher contagiousness.

**Figure 1:**
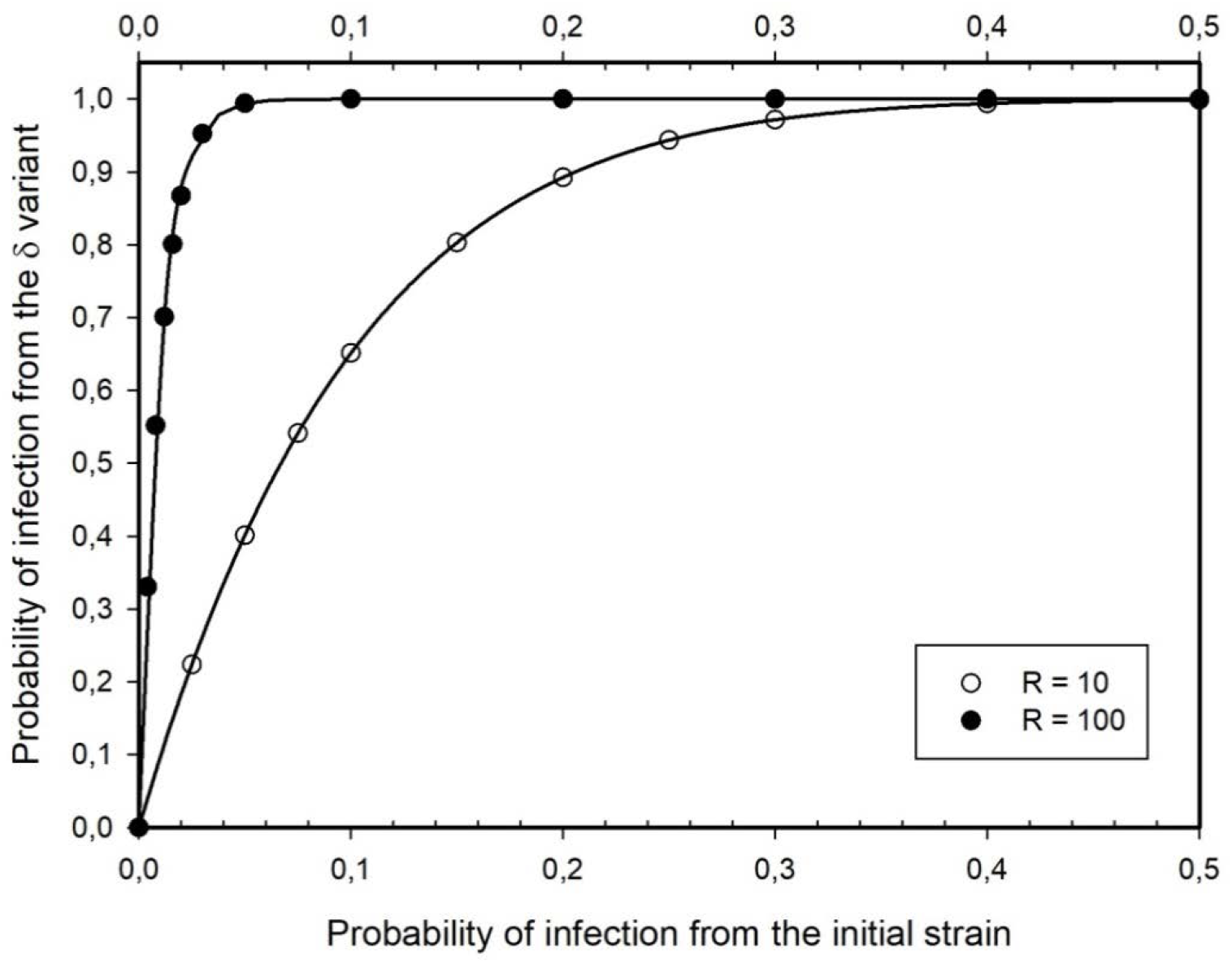
δ probability of airborne infection versus initial strain for a ratio of VL in respiratory fluids of 10 and 100 (all other parameters being equal).

### III-2. The case of public access area

We will examine first the case of an indoor space ventilated following the norm and at stationary state. Then, the dose of exposure is given by equation (10), and, in the Wells-Riley model, the probability of infection follows the Poisson law (11). If the ventilation of the public space *q*_2_ conforms to the norm per person *q*_*norm*_:

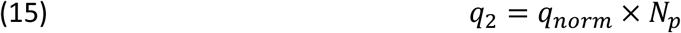

with *N*_*p*_ being the number of persons within the area. This assumption is of course questionable either if this norm is not followed or if the value of *q*_2_ is fixed constant, independently of *N*_*p*_ as it is often the case.

Assuming an infector proportion *r*, we can express the number of infectors as:

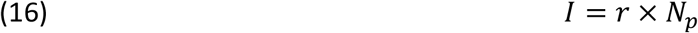

Strictly speaking it is the prevalence of infectors, including asymptomatic, that should be used for *r*. It is anyway probable that the number of infectors is proportional to *N*_*p*_. As discussed in SM4 it is extremely difficult to have the exact value of *r* from the values of positivity rate or incidence rate reported by health agencies. Below we use only “reasonable” values.

the dose of exposure results:

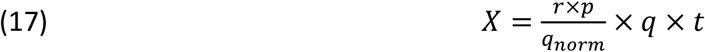

which clearly shows the multifactorial character of the risk. In the case where the ventilation conforms to the norm and for a given value of *r*, the difference between a school, a restaurant and a commercial center comes essentially from the time of exposure *t*. Note that this time is a total time which does not need to be continuous but can be a summation of hourly and daily exposition in the various spaces that the individual went through, due to the fact that the risk is essentially probabilistic. **<u>Clearly the difference in quantum production rate between δvariant and previous strain, plays an enormous role</u>** in the dose, and hence in the probability of infection. However, it is clear from equations (11) and (17) that the known parameters on which it is possible to play are the time of exposure *t*, the ventilation rate *q*_2_ itself, depending on the norm of ventilation *q*_*norm*_ and on the number of persons in the volume, if the total ventilation conforms to the norm.

Note that when *N*_*p*_ is not very high, Eq. (16) may lead to a number of infectors *I, I* < 1, which could seem unrealistic. Instead of the use of Eq. (17) for the dose used with the Poisson probability (hereafter *P*_*WR*_ – Eq. (11)) the following value of the probability should be used:

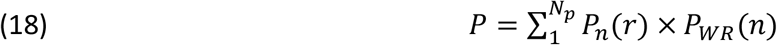

where *P*_*n*_(*r*) is the probability to have *n* infectors and *P*_*WR*_(*n*) the probability of being infected with *n* infectors.

Then, it can be shown, (see SM4) that equations (3) with (17) lead to a very similar result than the more exact calculation (18), assuming that the ventilation rate follows equation (15).

In Figure 2, the curves of equal probability of infection versus the time of exposure and the ventilation volumetric flow rate (starting at 5 m^3^/h/person) are shown, for a quantum production rate of 40 h^-1^, and an infector proportion *r* = 0.01. Of course, in the real life, if the ventilation rate is fixed at the maximum space occupancy and not by equation (15) it would result in a smaller probability of infection in a non-fully occupied room. Note that this figure results from the assumption that the ventilation rate is proportional to the number of people in the well-mixed space.

**Figure 2:**
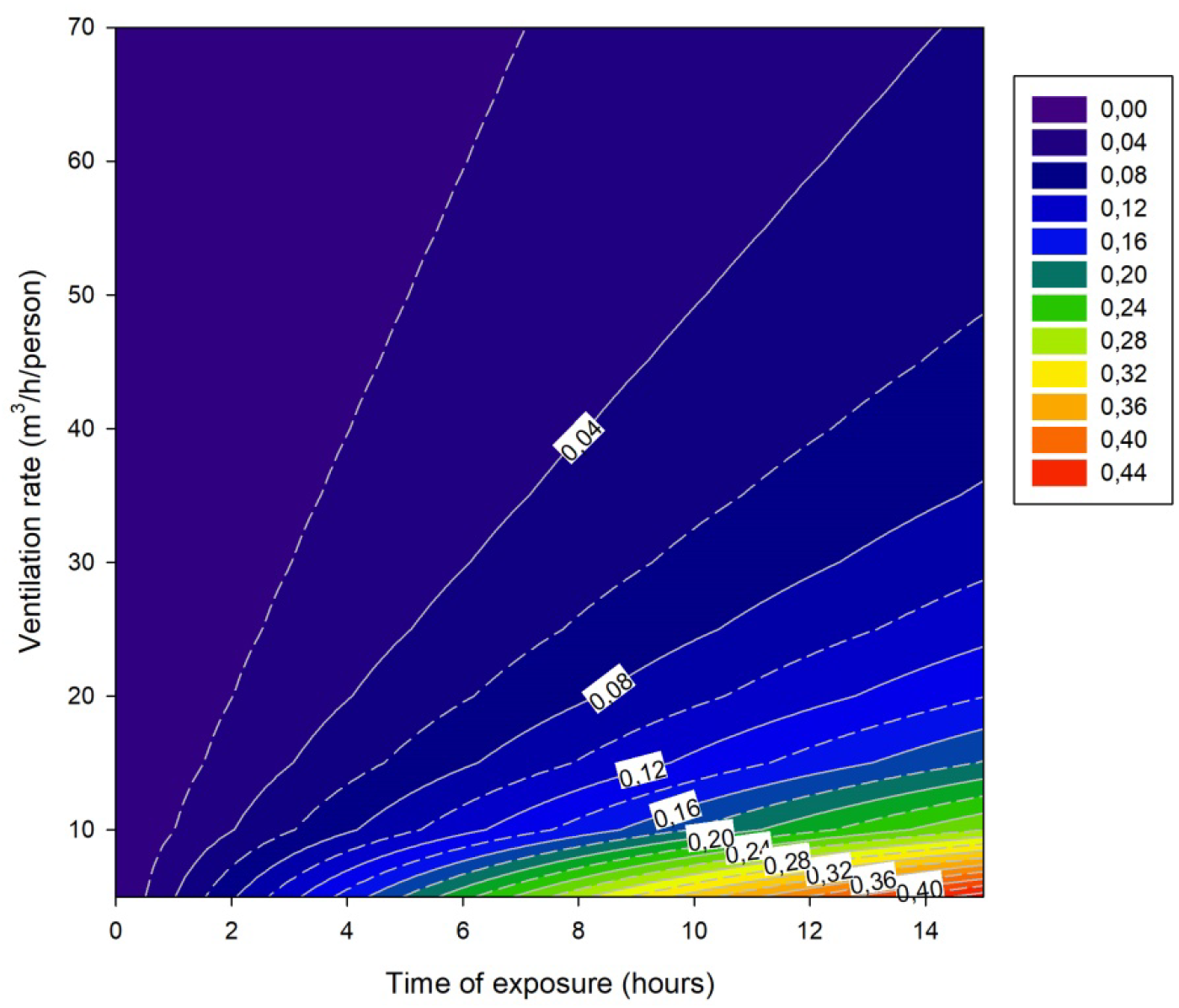
Probability of infection contours as a function of time of exposure and ventilation rate per person assuming a quantum rate of 40 h^-1^, an expiratory rate of 0.50 m^3^/h and an infector proportion of 0.01.

In the case of very poor ventilation, we can use Eqs. (3 and 12) in order to estimate the risk in a public space as a function of the number of persons in the volume V and of the time of exposure, assuming that at time *t* = 0 the concentration of quantum is zero. This could be for example the case of a poorly ventilated tutorial room (i.e. 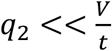) where the lecture (and hence the student presence) starts at *t* = 0; *t* being the time of exposure. Figure 3 displays the curves of equal probability of infection versus the time of exposure and the number of persons for an infector proportion of 0.01 and a volume of 150 m^3^.

**Figure 3:**
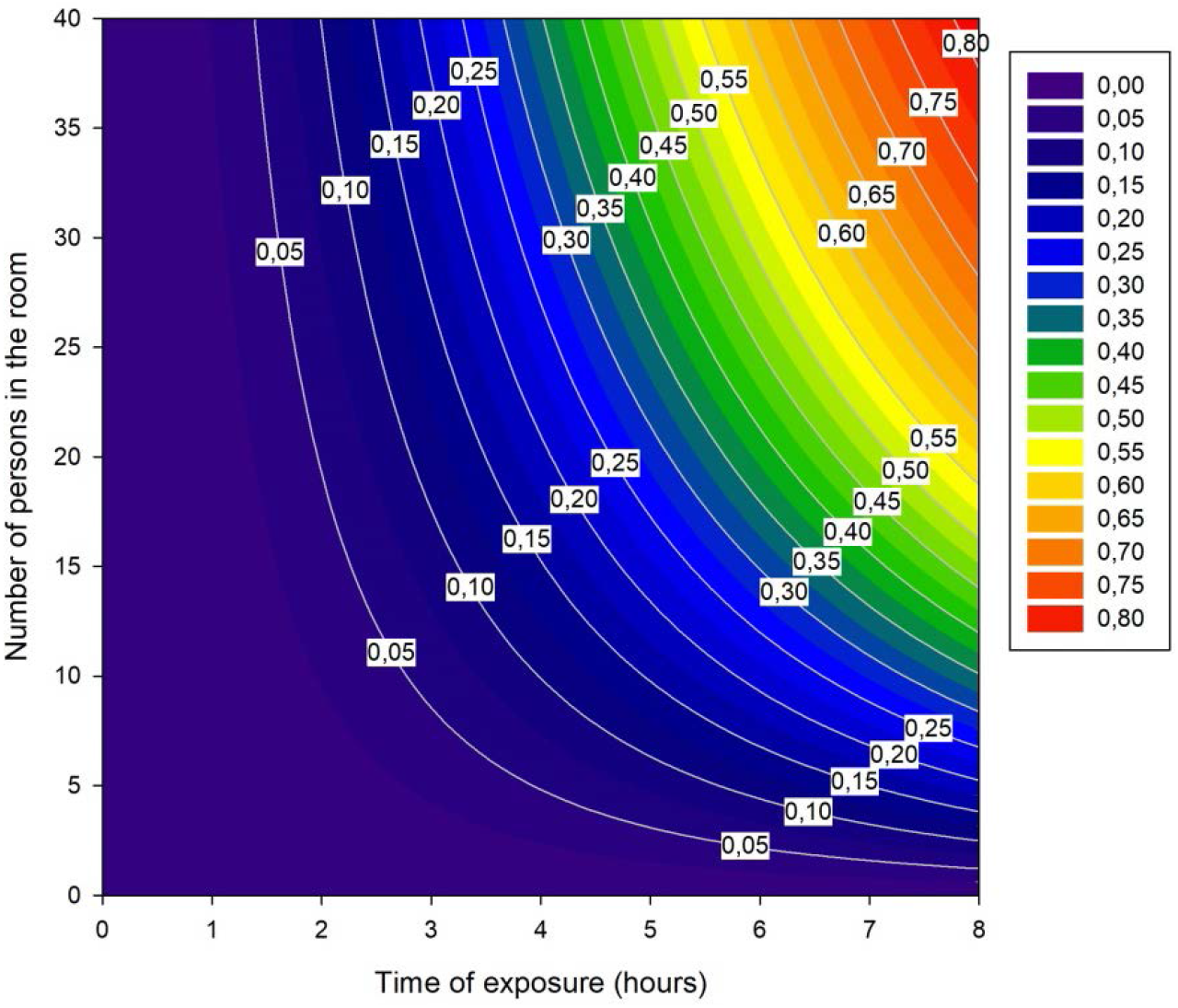
Probability of infection when the ventilation is poor (see II.5). Calculations are made using an expiratory rate of 0.50 m^3^/h; a quantum rate of 40 h^-1^; an infector proportion of 0.01 and a room volume of 150 m^3^ typical of a lecture room.

Note that the wearing of masks will of course alter these figures by reducing the quantum production rate as well as the quantum inhaled quantity (see SM5).

## IV Analysis of some specific cases

### IV-1. Observations

As discussed earlier, aerosols are the main contamination routes of COVID-19 and exposure becomes critical indoors. It is now widely admitted that ventilation is, beside the mask, the most effective way for reducing indoor airborne transmission [8,13,38,39] in particular for highly insulated and airtight buildings, where the building envelop infiltration is reduced to a minimum to respect thermic regulation. The measure of indoor CO_2_ concentration is considered in standards as an indirect measure of IAQ [7] or as a proxy of ventilation rate. One should distinguish the indoor CO_2_ limit values (1000 to 1300 ppm) issued from building ventilation regulations [40,41] from maxima recommended in the current sanitary context: 800 ppm wearing a mask and 600 ppm without a mask [42,43]. In fact, as recalled by Li [39], outside of healthcare settings, existing ventilation standards do not account for infection control. When CO_2_ concentration exceeds threshold values, the ventilation flow rates are usually insufficient and aerosol route contamination risk is high as illustrated by Figures 2 and 3.

In this context, we carried out, in autumn 2021, a series of CO_2_ concentration measurements and observations in various environments. Measurements consisted in determining the CO_2_ time evolution within each room using non-dispersive infrared (NDIR) CO_2_ sensors (Aranet 4 or ZG-106 Protronix CO_2_ monitor). Their accuracy was ±3% and ±5% of reading for the Aranet 4 and the ZG-106 Protronix respectively. The sensors are factory-calibrated and allow raw data logging with time stamps. Sensors were positioned between 1 and 2 m height (corresponding to the occupants head position), at least 2m far from every person and distant from windows or doors.

Further, when possible the mechanical ventilation was directly measured by using a balometer from ACIN (Flowfinder mk2). The accuracy in flow rate measurements was ±3% of the reading. Three categories of spaces were investigated including two university lecture rooms (ULR5 and ULR-20) and one pupil schoolroom; two university amphitheaters (UAW and UAE) and finally a restaurant. For each room, the main characteristics are given in Table 1. This includes, among others, the maximum allowed people from which the regulatory ventilation is determined according to French regulation [41] which specifies the flow rate per person (PFR hereafter) as being 18 m^3^/h/person for lecture rooms and amphitheaters; 15m^3^/h/person for the schoolroom and 22 m^3^/h/person for the restaurant. A time step of 10 minutes was sometimes fixed in accordance with the French IAQ decree n° 2012-14 [44] for five-days monitoring to determine the ICONE index (see SM6).

**Table 1:** Ventilation measurements for various environments with their own main characteristics

| Room | ULR5 | ULR20 | Schoolroom | UAW | UAE | Restaurant |
| --- | --- | --- | --- | --- | --- | --- |
| Volume (m<sup>3</sup>) | 136 | 402 | 173 | 900 | 1035 | -- |
| People/max | 28/30 | 67/68 | 30/30 | 40/142 | 95/163 | var./120 |
| measurement duration/time step (min) | 80/10 | 90/5 | 7days/10 | 56/ var. | 55/1 | 5days/5 |
| Ventilation system<sup>a</sup> | U | B-dyn | H | B | B | B or B-dyn |
| Regulatory volumetric flow rate (m<sup>3</sup>/h) | 540 | 1224 | 450 | 2556 | 2934 | 2640 |
| Volumetric flow rate from CO<sub>2</sub> (m<sup>3</sup>/h) | 53 | 1124/450 | 50-100 | 2576 | 1219 | -- |
| Measured volumetric flow rate (m<sup>3</sup>/h) | -- | Max/Min<br>=<br>1187/200 | -- | -- | 1009 | ~ 500 |

<sup>a</sup> B: bidirectional ventilation; B-dyn: bidirectional dynamic ventilation; U: unidirectional ventilation; H: hybrid ventilation

The CO_2_ time evolution followed the standard law:

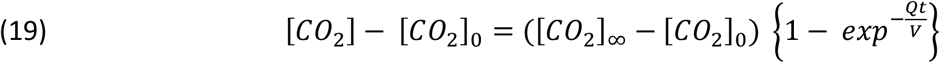

where [CO_2_]_0_ is the CO_2_ concentration, expressed in ppm, at the beginning of the analytical fit (*t* = 0), [CO_2_]_∞_ is the stationary CO_2_ concentration (*t* = ∞), *Q* the ventilation flow rate (m^3^/h), *V* the room volume and *t* the time at which the measurement was carried out. From this equation, it is straightforward to determine the ventilation flow rate *Q* from an exponential fit of the measurement when the volume *V* is known, at least when [CO_2_]_∞_ is not ill-defined, a situation that occurs when the number of people constantly changes with time like in the restaurant (see Table 1).

The CO_2_ time evolutions are illustrated in Figure 4-(a-d) where the reference of the CO_2_ concentration has been taken as an outdoor [CO_2_]_ext_ usual value of 400 ppm instead of considering [CO_2_]_0_ as the reference. This makes it easier for the readers to return to the absolute value since the initial [CO_2_]_0_ is never the same from one test to another.

**Figure 4:**
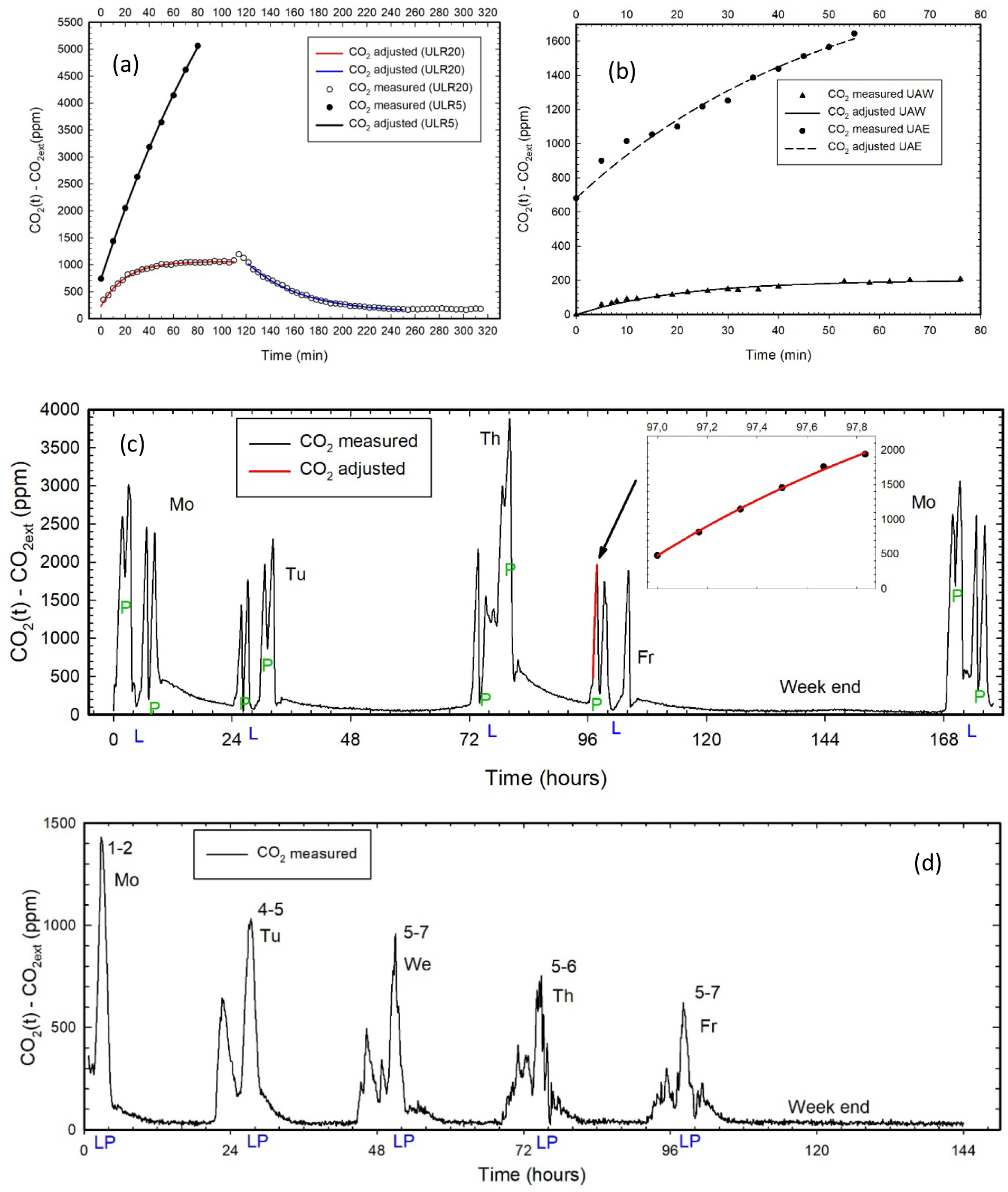
CO_2_ time evolution within examples of indoor spaces – complementary information are given in Table 1: (a) two lecture rooms (ULR5 and ULR20); (b) two lecture halls (UAE and UAW); (c) schoolroom over one week, L: Lunch, P: Playtime; (d) restaurant over a week (numbers close to the CO_2_ peaks represent the strength of the wind in Beaufort scale, LP: Lunch Peak).

Figure 4-a compares two lecture rooms (ULR5 and ULR20, see Table 1). These lecture rooms are at a University building over 50 years old, which has not yet undergone any energy retrofit. The ULR5 is equipped with air intake vents installed in window frames. As the building envelope is not airtight and since the toilets facilities, equipped with mechanical air exhaust, are far away from ULR5, little fresh air enters by the windows intake vents. In addition, exhaust flow rates at the level of the building are too low compared to the regulatory ventilation needs. This explains the observed very poor IAQ with maximum concentrations of CO_2_ exceeding 5000 ppm. This trend has been confirmed in a similar lecture room (ULR4, not shown for brevity) where CO_2_ concentration measurements during five consecutive scholar days lead to an air stuffiness index ICONE of 4, i.e. very high confinement (see SM6).

The ULR20 is a lecture room, among three rooms of the same previous building, which were fitted more than ten years ago with a common dynamic two-way ventilation system, using the level of CO_2_ in the exhaust circuit to control the ventilation flow rate. This system sized for a maximum flow rate of 1187 m^3^/h (for occupancy capacity of 68 students plus a teacher, i.e. 17.2 m^3^/h/person close to the French regulatory value of 18 m^3^/h/person). It is however set at a minimum flow of 200 m^3^/h during the unoccupied hours, and is manually switched off during holidays. In this room, on 2022/01/03, while the ventilation was still off after holidays, a maximum concentration of 3300 ppm was registered after one hour during an exam gathering 64 persons. The corresponding evolution is not given for brevity. During normal operation of the ventilation system of full occupied ULR20, the CO_2_ level does not exceed 1700 ppm (see Figure 4-a). This threshold corresponds to a Category 3 classification (moderate level may be used for existing buildings) in the UE regulation [42,45] and is above the French limit value of 1300 ppm [41]. However, this remains acceptable in comparison with the previous ULR5 case.

Figure 4-b presents CO_2_ evolutions in two lecture halls (UAE and UAW). UAE is, as previously, over 50 years old, whereas UAW is inside a modern new building. One can observe that in UAE the CO_2_ concentration reaches a high value of 2100 ppm after a one hour lecture gathering 95 persons. This corresponds to a PFR of 13 m^3^/h/person. Note however that when the lecture hall is full, the PFR would then be equal to 8 m^3^/h/pers., which is very far from the regulatory value. On the opposite, UAW seems very well ventilated since the CO_2_ concentration did not exceed 600 ppm in the presence of 40 persons. The deduced volumetric flow rate was as high as 2576 m^3^/h, which results in PFR = 18 m^3^/h/person when considering the UAW maximum capacity of 142. Therefore, this lecture hall complies with French regulations, and probably when it is full, the CO_2_ would be in the regulatory range 1000 to 1300 ppm [41]. However, we can regret, **for energy consumption reasons**, the apparent absence of flow rate control as a function of the occupancy density.

Figure 4-c shows the CO_2_ time evolution acquired during one full week in a classroom. The building is old (built almost a century ago) and has not benefited from any energy retrofit. The considered schoolroom receives 30 pupils 7-years-old. The insert in Figure 4-c gives an example of a CO_2_ rise from which the ventilation rate could be estimated. Since, the flow rate was found quite small, the measurements presented some dispersion from one day to the other but the observed range (50-100 m^3^/h) is very far below the regulatory flow rate for a schoolroom with a maximum occupancy of 30 persons (i.e. 450 m^3^/h according to the French regulation [41,46]). The corresponding air stuffiness index [44] is ICONE=4, corresponding to very confined class. This observation joins those of the French IAQ observatory [47] and various literature studies of ventilation state in schools in France [48,49] and elsewhere, particularly in Europe or USA [50]. In this latter investigation, Fisk performed a thorough review, which demonstrated the widespread failure of ventilation systems to provide the minimum flow rates specified in standards for classrooms. He reported that the maximum peak CO_2_ concentrations ranged from about 3000 to 6000 ppm. It is also important to stress that the French standard [41,46] makes the differentiation between young children (under 15 years old, PFR = 15 m^3^/h/person) and older teenagers or adults (older than 15 years, PFR = 18 m^3^/h/person), whereas this is not biologically relevant [51] because young children emit as much CO_2_ as older ones or adults. Children being more fragile than adults, the individual PFR should on the contrary be higher for them. The UE Regulation [42] recommends a PFR = 36 m^3^/h/person in the best IAQ category (category 1) for sensitive and fragile persons with special requirements, which should be the case for young pupils.

Finally, we carried out a CO_2_ monitoring during a week (Figure 4-d) in a modern restaurant situated in a coastal location of the Department of “Côtes d’Armor” in France. We used two Aranet sensors each one set in one of the two lunchrooms of the restaurant which communicate to each other through a large aperture. The two sensors were approximatively at a distance of 10 m to each other and demonstrate a similar CO_2_ concentration along the week. This is a strong demonstration that for this case, the well-mixed assumption holds. Interestingly, the restaurant is exposed to the wind, which can cause large variations in air renewal flow rates. Observations correlate strongly with an enhancement of ventilation with the strength of the wind (and inversely for CO_2_ concentration) which is shown on each peak of the figure in Beaufort scale (Bt = 1-2 on Monday; 4-5 on Tuesday; 5-7 on Wednesday; 5-6 on Thursday and 5-7 on Friday). Not indicated is the direction of the wind which has been changing continuously along the week. The high variability in peak CO_2_ from day to day can be clearly seen in Figure 4-d and wind effect on the level of airing appears obvious. On Monday, when there was no wind, a maximum concentration of 1800 ppm was recorded, which is a high level compared to the French public health committee recommendations to not exceeding 600 ppm in situations in which attendees are not wearing a mask [43].

Furthermore, in essence the restaurant is a place where conditions are continuously variable (customers do not arrive at the same time, doors open frequently) and it is not easy to establish stable conditions allowing to determine air flow rates from CO_2_ concentrations. Moreover, even if we do not have the confirmation, it is very likely that the bidirectional ventilation is dynamic, which makes air flow rates variable. The in-situ volumetric flow rate measurements done in customers’ space (lunchrooms and bar), lead to a total air flow rate around 500 m^3^/h. The hood in the kitchen and the related compensation grille, placed on opposite exterior wall, have probably an effect on flow patterns in lunchrooms, as the kitchen door is kept open during lunchtime. Since our objective was to evaluate the potential risk of contamination in a space where masks fall, we did not focus too much on a precise determination of the ventilation rate considering the above-mentioned difficulties. Rather we concentrated on the CO_2_ levels achieved every day (see discussion in section IV-2).

Through all above observations, the poor ventilation of the investigated premises is evident since most of our measurements range between one third and one tenth of the regulatory volumetric flow rates. Further to this failure in respecting norms, it is essential to understand that the present ventilation standards worldwide are not designed for infectious control, whatever the respiratory virus is. The present work also agrees with the large surveys of various bibliographical sources (Ribéron 2016, Canha 2016, Batiactu 2018) not only in France as revealed by the thorough review from Fisk [50]. Interestingly, in this latter study, Fisk mentions that increasing ventilation with annual costs ranging from a few dollars to ten dollars per person constitutes less than 0.1% of typical public spending on elementary and secondary education in the US. Such spending is judged a small price to pay given the evidence of health and performance benefits. This observation is more than ever true in this pandemic period and could be extended to other countries and other sectors than education. In the same spirit, it is desirable to generalize the use of CO_2_ sensors, a very affordable tool, in buildings to assist people in applying the suitable mitigation behaviours such as windows opening for instance to accelerate indoor air renewal.

### IV-2. Risk assessment

For the various situations described above it is important to derive a risk probability for an exposed person (susceptible) as a function of the observed CO_2_ concentration. From a statistical point of view and a large number of persons, the dose can be written (see SM7) as:

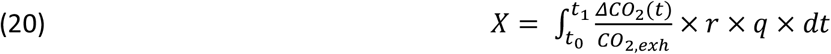

This relationship is valid for any situation including environments with poor ventilations and transient situations as well as stationary states 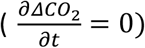. It does not need the ventilation flow rate value. It can be extended to include a virus lifetime (omitted here for sake of simplicity), which does not change the conclusions. Parameters *r* and *q* are again the proportion of infectors and the quantum production rate respectively, Δ*t* = *t*_1_ − *t*_0_ is the time of exposure of the susceptible, *CO*_2,*exh*_ the quantity of CO_2_ in the air exhaled by a human (∼40000 ppm), *ΔCO*_2_ the difference between the measured CO_2_ in ppm and the outdoor natural level measured with sensors.

We can define a mean value of “human” CO_2_ for the time of exposure Δ*t* by:

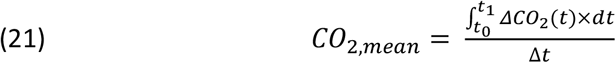

Then, the dose can be written:

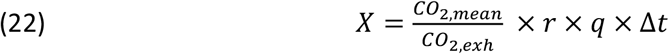

which highlights, beside the CO_2_ concentration, **<u>the importance of the time of exposure</u> Δ*<u>t</u>* <u>and of the number of infectors</u>**. Note that the remarks made in section III-2 for the *r* value remain valid. If a healthy subject is exposed to successive doses *X*_*i*_ corresponding to different periods of exposure Δ*t*_*i*_, then the total dose is just the sum of the successive doses (cumulative risk):

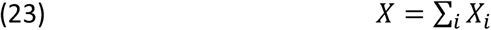

Following these formulas, we can deduce some risk probabilities corresponding respectively to the situations described in section IV-1. They are summarized in Table 2:

**Table 2:** Probability of infection for various scenarios

|  | school |  | restaurant |  | UAE | UAW | URL5 | URL20 |
| --- | --- | --- | --- | --- | --- | --- | --- | --- |
| $\Delta t(\text{min})$ | 1100 | | 80 | 400 | 55 | 76 | 80 | 112 |
|  | No mask | mask | 1 meal | 5 meals | 1 Lect | 1 Lect. | 1 Lect. | 1 Lect. |
| $P$ | 0,237 | 0,027 | 0,005<X<0,012 | 0,040 | 0,013 | 0,001 | 0,040 | 0,013 |

In this table we have utilized the same values for *r* and *q* as for figures 2 and 3: 0.01 and 40 h^-1^ respectively. Mostly for the values of calculated doses, the probability for a healthy susceptible to be infected is just nearly equal to the dose, due to its low value as explained previously. Therefore, the influence of changing the values of *r* and *q* can be easily estimated by a proportional calculation, as long as the dose remains small.

Some points in this table merit to be highlighted:

- For the school, the situation would be catastrophic without the risk reduction due to the mask. However, the precise quantitative impact of mask wearing is difficult to evaluate as discussed in the SM5. Also the social acceptability of mask wearing by children merits to be discussed.
- For the restaurant/bar, we have considered that customers are mainly workers who spend about 80 minutes at lunch. The risk is negligible for a single meal. In Table 2 *P* is bracketed since conditions varied depending on the day. If the restaurant is visited on a daily basis (5 meals) risk could be raised to a few percent following equation 23. However, the calculation does not consider that the mask is partly worn in the restaurant. In any case our observation and calculation show that the risk here is not especially high, which questions public policy in this field.
- For the other premises, which are located at the university, observations show a considerable dispersion. The risk can be very high for a lecture room very poorly ventilated (case URL5 of table 2) as well as reasonable in well ventilated area (case UAW). It must also be considered that for the university premises we have not considered either mask wearing or the cumulative aspect of the dose. As discussed in SM5, using masks induces a risk reduction of a factor of about 9. This is however easily counterbalanced within one week if students attend 9 lectures in the same room which is quite possible. It remains that in poorly ventilated areas the risk is high.

## V Implications of increased airborne contamination for health policy

The previous sections highlight the multiparameter character of the risk, through the time of exposure and the concentration of airborne infectious particles, itself linked to the proportion of infectors and to the indoor ventilation flow rate. With new variants such as δ or o (omicron), the quantum emission rate *q* can be estimated orders of magnitude higher than with the original strain due to VL or microbiological characteristics. Then, the spread of the virus should be mainly airborne even for close contact, and much more efficient. This increased spreading is in fact observed [52] even if, fortunately, it seems that the new variants are much less lethal than the original Wuhan strain. Moreover, health policies have not been sufficient to slow down efficiently this new contamination, especially in Western Europe. With more dangerous variants or new respiratory diseases, either more lethal or more contagious or both, new intervention measures must be considered. In the δ or o variant cases, the models and concepts presented in this paper and the experimental measures reported, lead us to derive implications **<u>for health policy</u>**. Such an exercise has already been done previously by leading scientists of the field [13] but it **seems that it has not been sufficiently considered** by health policies. Moreover, we do think that, beside a variety of engineering solutions already preconized by Morawska *et al*. [13] other mitigation measures are necessary, and we insist that authorities have to **change their mind in matter of priority**.

Amongst the various interventions of public policy discussed below we focus on the non-pharmaceutical ones. We first consider interventions directly targeting IAQ, i.e., mask, air filters and sterilizers, and ventilation. In this context we will also discuss the influence of the way of life, which depends on the country and the climate, and could lead to take immediate measures with strong positive consequences. We will then turn to interventions that are not directly targeting IAQ but nevertheless have implications on IAQ (e.g., living conditions during lockdown) or whose effectiveness is dependent of our understanding of contamination routes (e.g., contact tracing).

**<u>Such discussion is all the more needed</u>** that vaccine efficiency has been reported dropping far from 100% with time and variants for most vaccines, including Pfizer, and that their ability to stop transmission by asymptomatic infection is questionable [53,54]. Vaccination alone will not be enough to stop the epidemic spreading via airborne contamination, because present vaccines do not provide 100% immunity, especially with new variants such as omicron, although they result in a strong reduction of illness gravity. Beside the need of a **<u>large vaccination of people at risk</u>** (elderly, diabetic, overweight etc.) to reduce disease severity, it is clear that mitigation measures especially toward the problem of IAQ, should be highlighted: checking of HVAC (Heating, Ventilation, Air Conditioning) systems, air monitoring or development of high flux air sterilizers. New variants or new respiratory viruses in the future require a change of paradigm in this field [55]. If measures implying technological developments can be implemented only on mid-term, measures directed toward people information and the way of life must be taken immediately.

### V-1 Targeting IAQ

When IAQ is deficient, especially in indoor situation, **<u>wearing a mask is certainly highly useful</u>** [10,56,57], but their efficiency (especially for surgical ones) is not such that it could be the solution alone. It is possible to calculate that the risk probability *P* could be decreased by a factor of ten when both infectors and susceptibles wear it (see SM5). However, with new variants the quantum production rate increase could counterbalance this advantage. Moreover, in most countries, after a deny of mask interest, the choice of surgical ones in the general population has been made, although they are much less efficient than N95 respirators [58,59]. In some situations, the public should be informed of the better choice, depending on the IAQ (see sub-section V-2). People must be told that wearing mask under the nostrils is inefficient.

Therefore we conclude that **<u>wearing a mask alone, although useful, is insufficient to counterbalance the very high VL due to delta variant or the microbiological characteristics of omicron</u>**. Also social acceptability of masks on the long term is most doubtful. Therefore, we must take further corrective measures to improve IAQ.

IAQ has been recognized as a concern for public health and is addressed by building norms. However, IAQ policy has mainly considered the issue of chemical and particulate matter pollutants, excepted in the context of health care buildings, such as hospitals [60]. It is time to address the question of airborne pathogens “pollution” in the general population and its consequences for respiratory diseases. This will need a considerable change in the norms and recommendations for buildings (Meslem *et al*., in preparation), since, from this point of view, they are still in their infancy.

The problem is closely linked to building ventilation, which has been for centuries a natural ventilation, i.e., fresh air intake by voluntary or involuntary leaks on the building envelope allowing entrance and circulation of fresh air without real control. Since the first oil shock and the subsequent implementation of increasingly restrictive energy regulations, including today new constraints linked with environmental impact, things changed with buildings becoming more and more airtight and with HVAC technologies allowing ventilation control. The admission of fresh air is therefore minimized at the lowest value (hygienic flow rates) compatible with physicochemical IAQ, in order to save energy but frequently this leads to non-compliance with regulatory hygienic flow rates.

We recommend, in the context of new buildings and retrofit that is put in place, the in-situ **<u>verification of regulatory flow rates</u>**. This is often not done, because the regulations do not require it, as it is the case in France in the context of the regulation RT2012 [61]. It follows, as exemplified in this work, that **<u>introduced fresh airflows are much lower than the regulatory values</u>**. The ventilation professionals published an alarming report on the failure of the ventilation systems and demanded in 2018 that a certificate of receipt of these systems be delivered, like the certificate of receipt of airtightness of building envelopes mandatory in RT 2012 [62]. The next regulation RE2020 [63] applicable since January 2022 for residential buildings, takes a step forward by setting up an obligation to measure ventilation flow rates. However, one can object that this point is not subject to a building acceptance certificate. Another criticism is that verification of the airflows is not entrusted to an independent control office since ventilation system installers can make the flow rates measurements themselves. The Swedish experience of the OVK (Obligatory Ventilation Control) in place since 1991 [64] is shared in REHVA site [65] as an example to be followed by European countries and elsewhere. The Swedish regulation specifies that the first inspection of the ventilation system is mandatory when it is taken into operation. Then, regular inspections are mandatory every 3 or 6 years, depending on the building type (3 years interval for pre-schools, schools, and health-care buildings). Jan Sundell has fought for decades to put in place this OVK in Sweden, but he mentions in his last editorial letter [66] that it is not enough. HVAC engineers must be properly educated to the question of the IAQ, and its public health issues. He wrote in 2019 “*today in the United States or China, students are not taught properly about ventilation. They are taught to design air conditioning!!!”*

On the short term, either for natural or mechanical ventilations, increasing **<u>their flow rate should be achieved</u>** when possible. This could be done by slight opening of windows if necessary. In 2009, Nielsen [67] analyzed experimentally the transport process of particles and tracer gases and show that a high flow rate (i.e., an air change per hour ACH from 6 to 12 h^-1^) to the ventilated space reduces the level of viruses and bacteria in this space, without draught effect if sufficiently large supply areas are used. The increased energy cost has to be put in balance with the considerable cost (and economy impact) of present public policy in most countries. This is particularly true in public buildings.

In some cases like offices, classrooms, aircraft or cabins, where people stand mostly at the same desk/place, solutions as personalized, or piston ventilation [68], could be adopted. Computational Fluid Dynamics have shown recently that personalized ventilation performed the best to prevent cross-infection [69] compared to mixing ventilation, followed by displacement ventilation, impinging jet ventilation, stratum ventilation and wall attachment ventilation.

As discussed previously, sterilizing and filtering air has the same effect than fresh air ventilation. In his book of 1955 [4], Wells recommended a ventilation rate per pupil at school of 510 m^3^/h **which is an enormous value**, an order of magnitude higher than any current norm. Such flow rates imply an important energy consumption. Probably aware of this difficulty, Wells proposed a variety of solutions to sterilize air, and more particularly the use of UV lamps. Nowadays the insufficient ventilation of schools and nurseries is largely recognized [70].

In order to remove infectious particles of air, HEPA (High Efficiency Particulate Air) filters could be used. HEPA air filters can theoretically remove at least 99.97% of airborne particles with a size of 0.3 microns (μm). For efficient operation, the filters should be inspected quite regularly, and changed periodically. A clogged HEPA filter can have a large leak rate through the peripherical gasket [71]. The pressure drop through the filter can result in rather large energy consumption, beside the cost of system equipment and maintenance.

The COVID-19 crisis has led to a considerable development of air purifiers and sterilizers that use, amongst others, UV germicidal power, which is well documented for viruses [72]. It can be shown by calculation that the UV power required to efficiently sterilize large air flow rates is rather small [73]. Unfortunately, most of the sterilizer systems found on the market treat a much too low air flow rate. The reason is probably that generally this kind of apparatus includes functions such as VOC (Volatile Organic Compounds) treatment, and HEPA filters which results in higher costs.

Therefore, development of **<u>cheap air sterilization units of very high flux</u>** is clearly needed on the mid-term. It is worth noting however, that employing such devices will make CO_2_ diagnostics no more relevant since the proportionality of the active virus concentration to the CO_2_ one will not hold anymore.

The way of life itself has implications on the disease transmission. More than a year ago Rowe *et al*. made the prediction [74] that sub–Saharan Africa will not be stricken so much by the pandemic in the future due to airborne considerations. This low spreading of the disease has been observed up to now and a variety of explanations have been proposed [75,76]. Rowe *et al*. [6] have rationalized this observation considering an “outdoor” way of life in these countries, which includes housing without air conditioning (AC), with large natural ventilation to ensure refreshment and open outdoor markets instead of supermarkets. South Africa where the prevalence of AC is much higher has been more stricken and COVID-19 clusters have occurred there in closed supermarkets, most often equipped with AC [77].

Therefore, it can be thought that in many places of low latitude, like West Indies or Guyana, coming back as far as possible to the outdoor way of life could have immediate benefits. This necessitates **waiving of AC when possible and turning back to natural cooling**, which implies large current of fresh air. In many locations where heating cannot be avoided implying indoor way of life, besides increasing ventilation, outdoor activities (for example outdoor markets) should be encouraged.

The cheapest way to monitor pathogen IAQ is measurement of carbon dioxide concentration. Too often, The concentration level alone is used as a sufficient risk proxy, and a limit around 800 ppm has been proposed [78] as safe. We have shown throughout the present paper that communication on this limit is misleading, as it ignores completely the question of the **<u>time of exposure</u>**. We propose the development of **intelligent sensors** that could provide several integrated values of carbon dioxide concentrations. Time of exposure and mean concentration, as defined by equation (21), could be displayed by such sensors.

Last, close contact risk (except intimate i.e. < 0.6 m) is recognized as essentially airborne with again a key role of exposure time [37]. In many situations, contact between two individuals lasts less than fifteen minutes [79]. In this context, the risk drops to a very small value as soon as social distancing between individuals is higher than 1.5 m, correspondingly to the communication of government and health agencies. However, a misunderstanding of the real mode of transmission in this case has led to irrational measures such as organizing files in supermarket with obligation to use entrances different from exit. Although it has not been yet studied in the literature, staying in the wake of an infector in a file for several minutes is certainly riskier than crossing the infector. We recommend that, although social distancing must be encouraged, **<u>such measures</u>** directed against fast crossing **<u>should be removed</u>** since they are misleading for the public and could in fact induce higher airborne transmission.

### V-2 Implications for interventions that are not directly targeting IAQ

The most radical intervention to mitigate the pandemic has certainly been the various forms of lockdowns that, notably in western societies, constitute a major limitation to liberties and was unprecedented in non-war conditions. While first lockdowns might have been necessary, given the lack of governmental readiness to fight such pandemics in western societies, we now realize that, beyond the obvious socio-economic implications, it has a significant downside related to psychological isolation and mental health. Poorer families, children, women, and people experiencing mental disorders have been particularly harmed by lockdowns [80-82], and this measure should be taken in only the most extreme circumstances. Moreover, the efficiency of lockdown strategies is a matter of debates [83,84]. Deleterious effects on people and families who live in small apartments and closed places where IAQ is low is clear: **gathering people that have not been tested** in an indoor housing **for a long time could be very counter-productive**: it has been shown indeed that a large number of contaminations occur in family environment [85,86].

Far less radical, although very recent, contact-tracing apps on smartphones are the typical intervention that any digital policy would have considered to support health policy. Such apps were first introduced to help policy to fight the very lethal Ebola disease. However, their efficiency is dubious and their ethical character questionable. When air monitoring measures, discussed in previous sections, indicate a significant risk, the public should be informed in appropriate ways so that behaviors can be adjusted. For risk induced by aerosol-based transmission, intuitive and responsive user interfaces could be developed to visualize outbreak risks in various room of buildings and alert facility managers and users in a way that could be similar and complement that outbreak risks related to fomite-based transmission [87]. But mitigation measures such as contact tracing apps will have little effect against long range transmission by aerosols. These apps have not been designed to fight this transmission path of the pandemic and aerosol transmission was ignored at their inception. When aerosols are emitted from delta variant, it is the exhaled microdroplets concentrations in a given space that creates the major risk. Focusing on close crossing (less than one meter for more than 15 minutes as we did in France with stopCOVID) in a public space can be dangerous because people can feel safe (at least feel being well informed with their app), when in fact what they should be warned (possibly by their smartphone, but even better by public screens or specific systems) is about the situation over IAQ. Therefore, given the airborne danger of delta variant, we consider that contact tracing apps are inappropriate for at least three reasons: First, to be effective they require that a very large share of the population uses them for contact tracing which has been considered unrealistic [88] and is still the case. In fact, whereas contact tracing apps have been redesigned to be less intrusive (e.g Norway case) and their governmental communication to influence their adoption adapted in to be less coercive (e.g. France case), a common nudging tactic to influence their adoption has consisted in adding a number of features influencing individual benefits such as giving information about risky regions or allowing to show conformity to vaccination plans to access public places thus transforming a risk detection app into an information public health and a sanitary pass app. As a result, after vaccination campaigns, these apps have been hugely downloaded. However, the effective activation of the apps for personal risk detection is still very low. Second, as we emphasize in the present paper, relevant parameters, **<u>notably time of exposure to risk</u>**, and space, but not necessarily distance, to a likely infector, were not well understood by the project developers [89,90]. Typically, distance for technology such as bluetooth is critical for accuracy and reliability [89], but if the risk is related to the nearly homogeneous spread of virus in a given space, the issue is about detecting the level of risk in this space and not necessarily identifying the smartphone of the closest infector. Third, such apps may both develop bad habits in the population and creates another danger for increasing potential discrimination and problems [88] such as fear of mass surveillance as in Germany or Switzerland [91]. Conversely if major public spaces are equipped with air monitoring equipment – currently monitoring CO_2_ as proxy - that display public information about IAQ, contact tracing apps would not be needed. Such information would be permanently visible by the public on some fixed screens similar to clocks in such places. In addition, to increase their situational awareness [92], those who are or feel potentially at risk could check the safety of places on their smartphones by accessing a public website where measures of all displays would be available on a map with color indicators. Both of these solutions would require people being proactive. As the situation may vary a lot from place to place or evolve rapidly, the population at risk could also use a warning emergency system conveying alerts through a dedicated device [93] or some augmented reality app [87]. Given our experimentation measures, such public displays would be highly trustworthy thanks to their high representational fidelity (notably current, nearly exact and relevant (on these notions see [92])) of the CO_2_ measures and thus limit the use of such warning emergency systems to those at risk and not coerce all the population to acquiesce to a rampant form of data surveillance. The cost of such air monitoring equipment and public website will not be very high and they are a more ethical and scientifically valid choice, given the prevalence of the aerosol transmission path, than current digital policy based on smartphone close-contact tracing.

## VI Conclusion

The present health policies in many countries suffer from an original sin which was the deny of airborne transmission. The advent of strains such as δ or o leads to much higher quantum production rates, implying that spreading of the epidemic is now mainly airborne. However, the communication of most public authorities remains essentially directed toward avoiding close contact and fomites transmission. Even if the importance of ventilation and mask wearing is now acknowledged, strong decisions devoted to fight airborne transmission are not yet there. This is regrettable since some mitigations measures in this field will not negatively impact people life, as others such as lockdown.

Major implications for public health policies have not been drawn from the conclusion that new variants lead to dramatically airborne contamination. This is a significant conclusion of the present paper on which we draw attention. Following the approach of Wells [4,5] and of most recent researches ([36,37] amongst others) we derive simple formulas allowing to estimate risks in a variety of situations. Applied to some specific observations of CO_2_ level in a variety of environments they highlight the importance of **<u>the time of exposure</u>** in risky situations.

Another major contribution of this paper is to highlight several interventions that need to be introduced, modified, or could be suppressed. Some measures can be immediately taken at minor costs, such as increasing ventilation when heating and using natural cooling in hot countries, coupled to **<u>CO</u>**_<u>**2**</u>_ **<u>monitoring</u>** to bring back CO_2_ concentration to a satisfying level for the time of exposure.

We have shown that the ventilation systems, either natural or mechanical are often far of following norms that are already insufficient. Therefore, **<u>ventilation checking</u>** should be promoted, and **<u>norms need to be revised</u>** to include risk of pathogen transmission. Norms **<u>must include sterilizers</u>** able to recirculate large air flows and which need to be developed at a reasonable cost.

In the short term, even if these measures are costly, a first plan to implement them in places where public services are crucial such as hospitals and medical services [94,95], schools [96] is necessary [97]. **Notably it must be clearly communicated that risk is not only dependent on CO**_**2**_ **level, but also to the probability that an infector is or has been in the room and to the time of exposure**.

Finally, digital means should be directed at informing people (e.g. with appropriate screens or web applications possibly using augmented reality for particularly vulnerable persons, rather than digitally tracing their (social) behavior and surveilling them). With the introduction of smartphone-based contact-tracing apps further embedded in sanitary passes, the pandemic has considerably accelerated the pace of the transformation of western societies towards digital surveillance. While some initial intentions were hoped to be good, such trend is dangerous and shows that ethical use of the digital is still in its infancy.

We insist that thinking only in terms of social distancing or social interactions has become a paradigm that needs to be changed. Scientific literature demonstrates that we can be infected by close contact, but other situations can be dangerous due to the very nature of airborne transmission. As viruses can stay infectious in the air, we should not only consider the possibility of contamination in co-presence, typically when people face each other, but also when people follow each other in a file or even when infected people have left a poorly ventilated room. These scenarios need to be highlighted in public information.

And last but not least, when the present pandemic will be over, what will stay in the mid and long term is the necessity to **change our mind and norms in matter of IAQ**, in order to include this problem of airborne pathogen transmission, an enormous challenge for building technology.

## Data Availability

All data produced in the present work are contained in the manuscript

## Acknowledgements ?

“This research did not receive any specific grant from funding agencies in the public, commercial, or not-for-profit sectors.”

## Author contributions

BR initiated, conceptualized and led the study. BR, AC and AM did the measurements described in section IV and made the corresponding analysis. BR, AC, AM and FR contributed to the writing and reviewing of the paper.

## Supplementary materials

### SM1 Host entry characteristics

As discussed in the main paper the quantum of contagium, as defined by Wells [1], considers a variety of mechanisms including pathogen inhibition by host defenses. These defenses include, beside microbiological phenomena (immune response and others), some physical processes described below that are important for contamination by the aerosol route.

In a series of remarkable experiments with rabbits and mice, Wells demonstrated that, concerning aerosols, very fine particles (which include dry nuclei) have a much higher infectious power than coarse particle, at least for disease such as tuberculosis and influenza. Wells’ explanation was that the human body has a very efficient system to prevent coarse particle larger than a few micrometers to penetrate deep in the respiratory system. Beside defenses against very coarse particles, specific to the upper respiratory tract (nostrils, nasal cavity, mouth, throat, pharynx), and voice box (larynx)), mucociliary clearance is a primary innate defense mechanism of the lung (see the reviews by Bustamante-Marin and Ostrowski [2] and Kuek [3]) that helps to remove smaller particles and pathogens from the lower respiratory tract, using the epithelium formed by ciliated and secretory cells. These later provide a mucus which is expelled by cilia toward the digestive system after swallowing. It is known that most respirable pathogens do not provoke illness when ingested, and there is currently no evidence that COVID-19 could be transmitted by ingestion [4]. Note that the mechanism of very fine particles deposition into the lungs has been the subject of numerous studies for mineral toxic dusts, such as asbestos [5].

Nowadays, the formidable progress of microbiology allows studying the influence of cellular characteristics on the vulnerability of cells to coronaviruses, which start with binding of the viral spike (S) proteins to cellular receptors [6]. Following some data, it has been anticipated that infectivity was higher in the upper respiratory tract and that the nose was a primary target [7]. However the severity of the COVID-19 is linked to the occurrence of pneumonia, followed by acute diffuse alveolar damage, which can be due to direct lung infection by airborne microparticles [8,9] or by indirect infection from the oropharynx to the lung by aspiration of the viral inoculum when breathing [7]. Also the study of nonhuman primate model reveals, after autopsy, the importance of lung lesions in macaques [10]. It seems reasonable to assume that, when the virus reaches the lungs directly, before some immunity able to inhibit viral reproduction has been acquired, it could result in devastating pneumonia, as sometimes reported in young, healthy subjects.

It has to be noticed that as well the remarkable experimental results of Wells for particle size than the most recent findings of microbiology cannot be directly used to develop a quantitative model of transmission risk. Therefore, some concepts and approaches must be developed prior to the establishment of any risk model.

### SM2 Conservation and transport equations

It is far beyond the possibility of this section of the supplementary materials to develop the complexity of transport and conservation equations for diphasic turbulent fluids, with the target of precise calculations of the fields of velocities, temperature and concentrations of the various components. We shall just present the equation used in the main paper for the case of a well-mixed room (homogeneous hypothesis) and the approach underlying much of the calculations used in inhomogeneous models in order to calculate the concentration field of infectious particles.

In a homogeneous model, it is assumed that there is no spatial gradient of risk in a space where the infectors and the receivers either evolve or stay in place. In other words, it is assumed that the infectious microdroplets are evenly distributed. This is typical of two kinds of situations. It happens first instantly in a space where high performance mixing ventilation is achieved using special air terminal units designed to promote a high jet induction (i.e., vortex diffusers, lobed diffusers). This case lies to forced convection state. In absence of this kind of mixing ventilation there are a variety of air motions induced by other phenomena, such as natural convection, wake of moving people, door openings for letting people in or out. It can be shown that in many situations of this sort, the well mixed room hypothesis is also valid [11]. Then, we consider an evenly distribution of microdroplets obtained by induced turbulent flows, although this distribution is not really continuous due to its discrete character (very low concentration). Using CO_2_ as a proxy of infectious microdroplets (i.e. quanta), observations show that this condition is most often fulfilled (see main paper).

Of course, if specific ventilation techniques are used [12], the generated directional air flows within the room lead to preferential aerosols trajectories following air distribution patterns.

In a homogeneous model it is possible to write a conservation equation for the concentration *n*_*i*_ of mono-sized microdroplets in a volume *V*, as developed by Rowe et al [13] for the indoor risk assessment, to compare with the outdoor case. Figure SM2-1 depicts the situation:

**Figure SM2-1:**
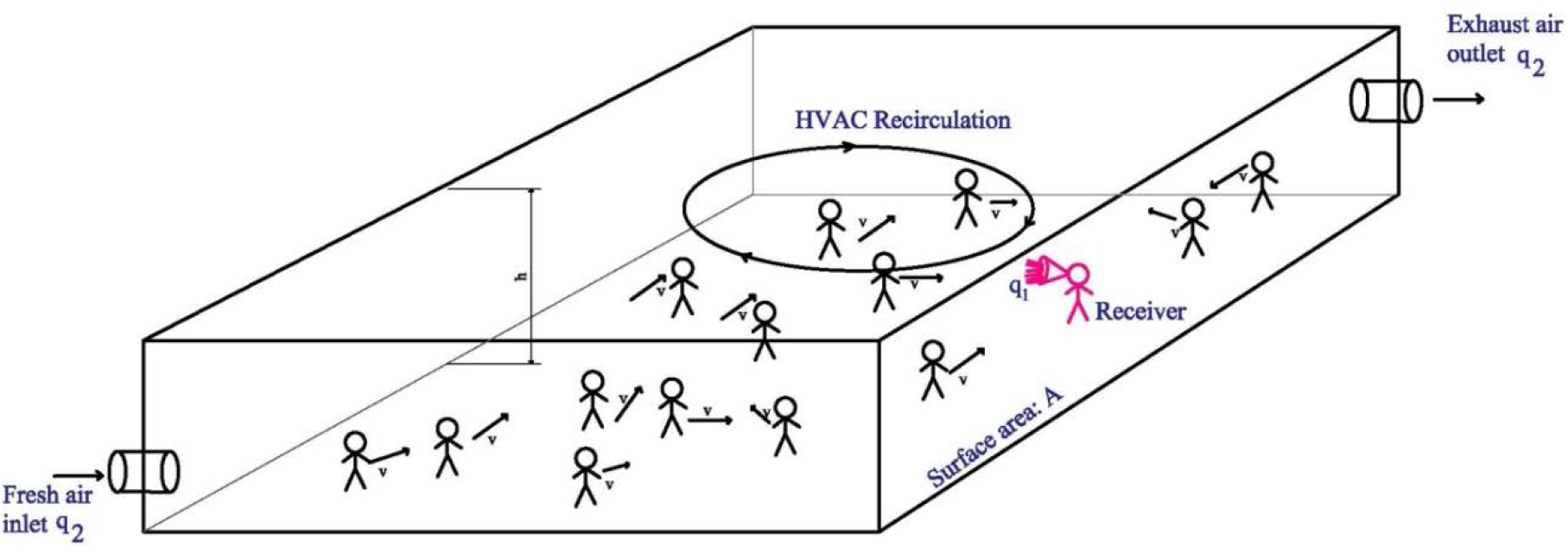
a typical indoor homogeneous situation.

In this figure the inlet and outlet ventilation flow rates are assumed equal with the value *q*_2_. Let *N*_*p*_ be the number of people inside, *N*_*i*_(*t*) the total number of aerosol particles of human respiratory origin inside the volume, resulting in a concentration of particles of *n*_*i*_(*t*) = *N*_*i*_(*t*)/*V*. The mean exhaled flow rate of a person was taken as *p* (of course identical to the inhaled rate) and the concentration of particles in this flow was assumed equal to *n*_1_

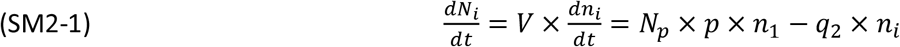

It was assumed no sink term for the particles inside the volume.

In the same way, an equation of conservation can be applied to the quanta of contagium as defined by Wells [1]. Let *N*_*q*_ be the total number of quanta in the volume *V* and *n*_*q*_ the quantum concentration. Considering the quantum production rate per infector *q* and introducing a quantum lifetime, which can be considered as the virus lifetime, *τ*_*i*_, this equation reads:

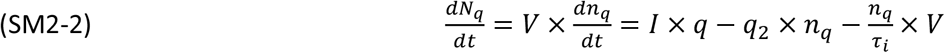

In this equation we consider the number of infectors *I* within the volume since only infectors emit quanta.

Assuming *n*_*q*_(0) = 0, The solution of (SM2-2) is:

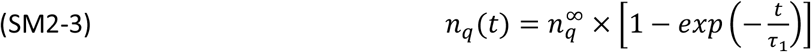

with:

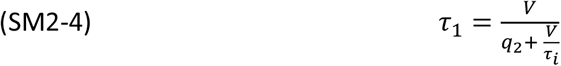

The concentration of quanta at stationary state i.e. *t*∼*a few τ*_1_ is:

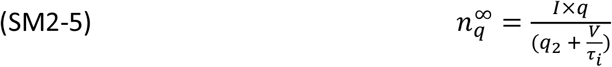

which, if the virus lifetime is neglected, reduces to:

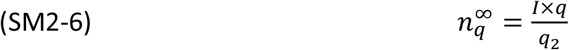

Note that if a device able to sterilize a flow rate *q*_3_ is used, the above equations hold just by replacing *q*_2_ by *Q* = *q*_2_ + *q*_3_.

These equations funded on the well mixed room hypothesis are the basis of the famous Wells-Riley model and are convenient for a very large number of indoor situations. However, inhomogeneous infection patterns are reported for a number of well-documented transmission events in closed spaces, especially in restaurants [14-16] but also in other places such as aircrafts [17]. Generally, in these specific well studied cases, inhomogeneity was created by the mechanical ventilation system of air conditioning (hereafter AC) with recirculation, inducing locally larger air velocity. One typical and largely mediatized event concerned a restaurant in Guangzhou, China. It has been the subject of numerical modeling [14]. Numerous published works in the field do not relate to a specific observed event but to hypothetical situations supposed to represent typical cases, such as a supermarket [18]. These models rely on CFD (Computational Fluid Dynamics) calculations of the air flow stream, using a variety of software, such as Open Foam for example. Then the microparticle behavior is estimated using a variety of methods (Lagrangian, Monte-Carlo). In the Lagrangian approach the movement of each particle is calculated using Newton’s second law of motion, where, within forces acting on the particle, the drag one is determined from the calculated field of air velocity. Note that, for a Stokes number << 1, the particles are just assumed to follow the flow. The Stokes number can be defined as the ratio of two times *τ*_*a*_/*τ*_*h*_, *τ*_*a*_ being the time of velocity accommodation of a particle to the flow velocity and *τ*_*h*_ the hydrodynamic time (equal to a typical length of the problem divided by the flow velocity). The Stokes number reads [19]:

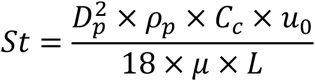

With *D*_*p*_ and *ρ*_*p*_ being respectively the diameter and volume mass of the particle, *μ* the air viscosity, *u*_0_ and *L* respectively a typical order of magnitude of flow velocity and length. *C*_*c*_ is a slip parameter which takes into account the value of the particle Knudsen number. For particles of the size considered in airborne transmission *C*_*c*_ is very close to one. Note that *L*/*u*_0_ is the hydrodynamic time and that for most problems dealing with the behavior of exhaled aerosol particles in indoor situation the Stokes number remains much smaller than one, except for large particles in the close contact case discussed in next section.

Il is also worthwhile to point out that when inhomogeneous infection modeling is applied to a specific geometry of the environment, it can be applied as such for the design of a new building for example but is limited for applications in the real life of most existing buildings and therefore, on the short term, for driving public policy. What is more interesting is the modeling of airborne close contact discussed in the next section.

### SM3 Close contact transmissions

It is now largely admitted that the transmission of COVID-19 disease by close contact is most often an airborne one, referred in the literature as “short-range airborne transmission” [20,20,21,21]. Close to the emitter the turbulent expiratory plume (or puff for cough and sneeze) can have a much higher quantum (viral) load than in the ambient air of the indoor space considered. Several models of this phenomena have been proposed, some very simple [20] others more sophisticated. The recent one by Cortellessa *et al*. [21] employs CFD for the air flow and Lagrangian calculations for the particles to derive the dose and the risk as a function of the distance between infector and susceptible. Not only the distance but also the time of exposure is considered in order to assess the risk, although the time is limited to fifteen minutes. Large microdroplets which behave in a ballistic way are also considered and shown to prevail only at very short distance (< 60 cm), with a contribution to the dose being completely negligible at higher distances, demonstrating the airborne character of most airborne contamination in close contact, excepted intimate.

In their paper, Cortellessa *et al*. also made a comparison with the homogeneous risk. However, the comparison is restricted to the same time of exposure of fifteen minutes, with an initial concentration of quanta equal to zero. Therefore, it does not consider long times of exposure for the homogeneous case at steady state, as found for example in schools but such an extension can easily be done. Indeed, a good comparison should have to include the probability of close contacts together with contact durations, which is not done. Such a close contact risk assessment is anyway extremely useful for public policy.

### SM4 Infector proportion and combination analysis

The problem of determining the exact proportion *r* of infectors *I* in a large population *N*_*Tot*_ (*r* = *I*/*N*_*Tot*_) is a difficult one. Two statistical results are most often available. The positivity rate is the number of populations tested positive related to the total number of people tested, and therefore is a proportion without dimension. The incidence rate is the number of new people tested positive in a population, which can then be reported to a target population (for example 10^5^ individuals) for a given period of time (for example one day or one week). It is therefore a temporal rate and, as such, has the dimension of (time)^-1^. It is clear from these definitions that the results will depend on which people are tested and also of the size of the target. Since many people are infected but not tested and that people tested positive in the past remain infectious for some time, it can be anticipated that the real number of infectors could be much higher than what can be deduced from an analysis of the incidence rate: in principle, this rate can drop to zero with still infectors in the population. Further, since the population tested is often a symptomatic one, the positivity rate of testing could be much higher than the real proportion of infectors. Only a blind testing of a representative population would lead a true value of *r*.

Therefore the purpose of the present SM is just to show that it is possible to estimate the probability of infection of a susceptible target using a simplified expression (see SM4-3) which essentially considers the given proportion of infectors *r* in a population of *N*_*Tot*_ individuals, provided that the ventilation flow rate per person, *q*_*norm*_, is known and the time of exposure *t* is fixed. Here *N*_*Tot*_ will represent the inhabitants of a country, a region, a metropole or a city or it can also denote a fixed reference population like 100000, for instance. Then, *N*_*Tot*_ is large. The number of infected people in that population will be quoted *I* further in the text (see SM4-6 and beyond) with *I* = *r* ×*N*_*Tot*_.

In the main paper we have derived an equation for the dose inhaled by a susceptible person:

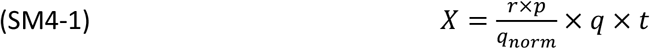

which assumes that the total ventilation rate *q*_2_ is given by *q*_*norm*_ × *N*_*p*_, where *N*_*p*_ is the number of people present, with the susceptible target, in a specific location. Here *N*_*p*_ « *N*_*Tot*_. It is also assumed that the proportion of infected people *r* is also representative of the sanitary situation in the space of interest. In other words, if *n* is the number of infectors in the restricted population of *N*_*p*_ persons, we assume that *r* = *n*/*N*_*p*_ = *I* /*N*_*Tot*_. We also remember that *p* and *q* are the respiratory flow rate and the quantum rate of pathogens per infector expressed in h^-1^, respectively.

From this, the probability of infection is given by the Wells-Riley expression already presented in the main text (eq. 3):

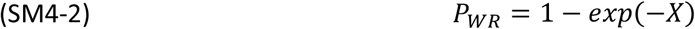

or

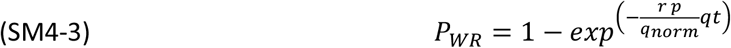

Another way to calculate this probability, which seems to be more realistic, is to make a weighted summation of probabilities to be infected in conditions where one, two, three etc. infectors are present in the restricted population of *N*_*p*_ people. This can be expressed as:

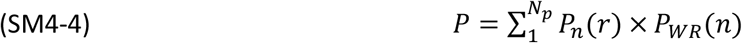

where *P*_*n*_(*r*) is the probability to have *n* infectors and *P*_*WR*_ (*n*) the Wells Riley probability of being infected with *n* infectors in the population of *N*_*p*_ individuals. Then:

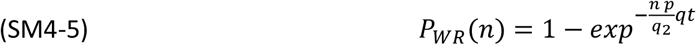

This new expression SM4-4 has an interesting advantage with respect to the simpler equation SM4-3 since it discriminates the individual *P*_*n*_(*r*) contributions from each other. Then, it is possible to evaluate how significant is each term in the summation and more particularly if the state with only one infector can be representative of the total risk of infection or not.

Probability *P*_*n*_(*r*) is dependent on the number of infected people *I* and consequently it is also a function of *r*. It can be derived from a combinatory analysis. Defining 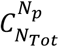 as the number of combinations of selecting an ensemble of *N*_*p*_ persons in a larger group of *N*_*Tot*_ individuals, one can express the number of combinations that include *n* individuals with a given property (here infection) in the selected group of *N*_*p*_ people. Then the probability of having *n* individuals infected in the restricted population *N*_*p*_ is simply given by:

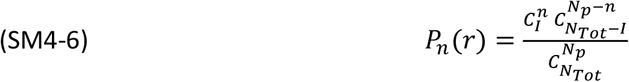

We remember here that the number of combinations of *i* elements in a global ensemble of *j* objects (with j ≥i) is mathematically equal to:

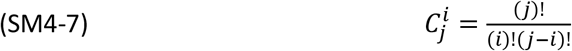

From this, equation SM4-4 becomes:

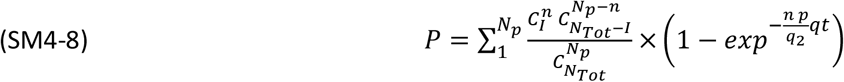

This expression is numerically evaluated below for a few examples and compared to equation SM4-3. We consider here situations for which the restricted population is smaller than the total number of infectors in the reference population *N*_*Tot*_:

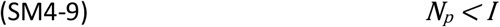

Calculations are be made considering a reference population *N*_*Tot*_ of 10^5^; a respiratory flow rate *p* of 0.5 m^3^/h; a quantum infection rate *q* of 40 h^-1^ and a time of exposure *t* of 2 hours. A standard ventilation flow rate *q*_*norm*_ of 20 m^3^/h/person will be also employed. The proportion of infected people *r* is varied between 0.001 and 0.03 and the restricted population *N*_*p*_ is chosen as either 80 or 30. From this, the number of infected people in the *N*_*Tot*_ main group will vary from 100 to 3000 according to the *r* value, thus respecting inequality SM4-9.

Results of SM4-3 and SM4-8 are presented in Table SM4-1 and SM4-2 for the two values of *N*_*p*_. In addition, we indicate the limit of *n*, quoted *n*_*cut*_, beyond which *P*_*n*_(*r*) × *P*_*WR*_(*n*) terms do not contribute significantly to the summation in SM4-8; the value of *n*, quoted *n*_*max*_, corresponding to the main contribution 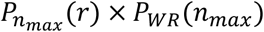 in the summation and the percentage of this contribution to *P* value.

**Table SM4-1:** Comparison of *P*_*WR*_ with *P* for a restricted population *N*_*p*_ of 80 individuals

| $r$ | $P_{WR}$ (SM4-3) % | $P$ (SM4-8) % | $n_{cut}$ | $n_{max}$ | $n_{max}$<br>contribution % |
| --- | --- | --- | --- | --- | --- |
| 0.001 | 0.200 | 0.197 | 2 | 1 | 93 |
| 0.003 | 0.598 | 0.591 | 3 | 1 | 79 |
| 0.010 | 1.980 | 1.956 | 4 | 1 | 46 |
| 0.030 | 5.824 | 5.756 | 8 | 3 | 27 |

**Table SM4-2:** Comparison of *P*_*WR*_ with *P* for a restricted population *N*_*p*_ of 30 individuals

| $r$ | $P_{WR}$ (SM4-3) % | $P$ (SM4-8) % | $n_{cut}$ | $n_{max}$ | $n_{max}$<br>contribution % |
| --- | --- | --- | --- | --- | --- |
| 0.001 | 0.200 | 0.193 | 2 | 1 | 97 |
| 0.003 | 0.598 | 0.579 | 2 | 1 | 92 |
| 0.010 | 1.980 | 1.917 | 3 | 1 | 75 |
| 0.030 | 5.824 | 5.644 | 5 | 1 | 43 |

These calculations demonstrate a very good agreement between both ways of determining the probability of infection from either *P*_*WR*_ or *P*. The agreement is even better when the restricted population is enhanced, essentially due to the statistical effect of using larger *N*_*p*_ numbers. It can also be shown that the contribution of one infector (*n*_*max*_=1) in the summation is the main one in many situations although, however, summation cannot be limited to the first term in SM4-8 for several conditions as indicated by the *n*_*cut*_ value and the “*n*_*max*_ contribution” columns. The lower the proportion of infectors *r*, the larger the contribution of *P*_1_(*r*) × *P*_*WR*_ (1) which makes a lot of sense since for small *r* the probability of having more than one infector in the restricted population *N*_*p*_ becomes very small.

To conclude we stress that we have restricted the demonstration to a limited number of configurations but it is worth pointing out that several parameters act in a similar way mathematically speaking. Then, changing the time of exposure or/and the quantum rate of infectors would lead to essentially the same kind of conclusions.

### SM5 Masks, quantum production rate and inhaled dose

To build a probabilistic model of infection it is necessary to know the production rate of quanta (as defined by Wells) by an infector. It is defined per unit time and per infector (unit: h^-1^ for example) and can be deduced from epidemiological observations [22] but also linked to the distributions of microdroplets emitted by humans, together with the knowledge of viral load in respiratory fluids and of the mean number of viruses required to infect 63% of susceptibles.

As stated in the main paper and following Buonanno *et al*. [23], the quantum production rate *q* can be written as:

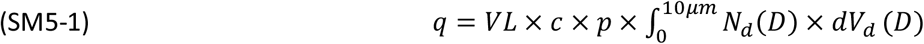

where *VL* is the viral load in the respiratory fluid, *c* is a factor of proportionality between the viral content (copies/unit volume) and quanta, *p* is the pulmonary exhaled volume rate (volume/unit time), *N*_*d*_(*D*) the size distribution of droplets (diameter *D*) of volume *V*_*d*_.

Morawska *et al*. [24] have shown that microdroplets emitted by different expiratory activity correspond to four different modes of size distribution, centered on mid-point diameters of respectively *D*_1_ = 0.8, *D*_2_ = 1.8, *D*_3_ = 3.5, and *D*_4_ = 5.5 μm. Their concentrations depend on the expiratory activity as shown in table SM5-1 adapted from Table 1 of Buonanno *et al*. [23]:

**Table SM5-1:** Concentrations (in cm^-3^) of the microdroplets size modes during various expiratory activities

| Expiratory activity | Centered mid-point diameter ($\mu m$) | | | |
| --- | --- | --- | --- | --- |
|  | 0.80 | 1.8 | 3.5 | 5.5 |
| Voiced counting | 0.236 | 0.068 | 0.007 | 0.011 |
| Whispered counting | 0.110 | 0.014 | 0.004 | 0.002 |
| Unmodulated vocalization | 0.751 | 0.139 | 0.139 | 0.059 |
| Breathing | 0.084 | 0.009 | 0.003 | 0.002 |

It results that equation (SM5-1) can be simplified as:

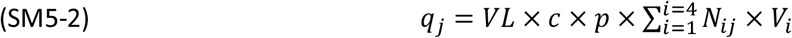

where the subscripts i and j refer to the size mode and the expiratory activity respectively.

From equation SM5-2 and Table SM5-1 it is clear that the production rate of quanta can vary widely depending on the expiratory activity but also on the virus strain through *VL* and *c*. Note also that the level of activity (which implies a given metabolism) plays an important role on this rate [23]. Therefore, it can change with time for a given infector.

For a given respiratory activity, equation (SM5-2) can be written as:

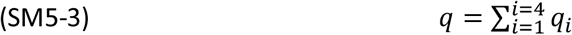

where the subscript j has been omitted.

In the absence of masks for the emitter (infector) and the receiver (susceptible) the dose inhaled by the receiver can be written:

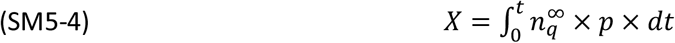

where 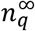 is given by equation SM2-6.

When a mask is worn the proportion of particles going through the mask could be strongly dependent of the particle size. Therefore, it could be considered that the quantum production rate is reduced accordingly and that it is possible to define a quantum production rate depending on the mode:

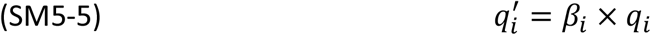

As a conservation equation can be written for each mode, a concentration of quantum for this mode at stationary state will result:

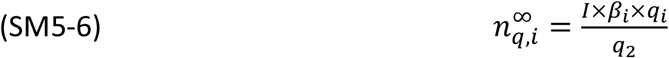

If the receiver wears the same kind of masks the inhaled dose of this mode of particles should be:

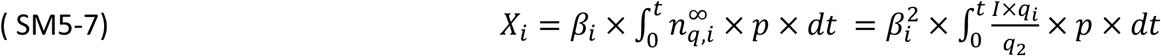

Then the total dose would be:

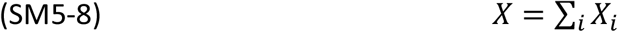

For the smallest size, below 1 μm (*i* = 1), the surgical mask can be very inefficient as shown by [25,26] leading to a value of *β*_*i*_ close to 0.5 for the flow through the filtration media.

However due to the importance of the leaks [27], it could be assumed that *β*_*i*_ is also very large even for particles larger than 1 μm (except for the largest ones which behave in a ballistic way and are completely trapped). Then, using equations SM5-7 and SM5-8 with the results of [26,27], it can be shown that wearing the mask reduces the quantum production rate by a factor of three. As the dose of inhaled particles is reduced by the same factor, an overall efficiency in dose reduction of around 90% can be assumed if emitters and receivers wear it, as it has been assumed for schools in the main paper.

### SM6 The ICONE index

Based on indoor CO_2_ concentrations, the ICONE air stuffiness index [28] has been developed in 2008 by the French Scientific and Technical Center of Building (CSTB) especially for IAQ evaluation in schools. In 2012, the ICONE air stuffiness index has been integrated into the framework for the mandatory monitoring of IAQ in some public buildings in France (IAQ decree n° 2012-14 [29]. The ICONE index takes into account the frequency and intensity of CO_2_ levels around the threshold values of 1000 and 1700 ppm during normal occupancy of the classroom by children. The confinement level is then expressed by a score scaled in six levels from 0 to 5. The score 0 corresponds to zero confinement (CO_2_ level always below 1000 ppm), this is the most favourable situation. Notes 2 and 3 correspond to low and regular confinement, whereas notes 4 and 5 correspond to very high and extreme confinement, level 5 is the most unfavourable situation (CO_2_ concentration always above 1700 ppm during occupancy). In this case, the decree [29] stipulates that additional investigations must be carried out and the local authority (the departmental Prefect) must be informed. Table below summarizes the various situations:

| ICONE | Stuffiness level |
| --- | --- |
| 0 | None |
| 1 | Weak |
| 2 | Moderate |
| 3 | High |
| 4 | Very high |
| 5 | extreme |

The icone index can be calculated precisely using the following expression:

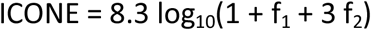

where f_1_ and f_2_ represent the proportions of CO_2_ concentration measurements comprised in between 1000 and 1700 ppm or higher than 1700 ppm respectively. Hence, the ICONE index is zero when all measurements have been found below 1000 ppm (f_1_ = f_2_ = 0) as said earlier whereas it is 5 when all measurements are higher than 1700 ppm (f_1_ = 0 and f_2_ = 1).

### SM7 The concentration of carbon dioxide as a proxy of the quantum concentration

The exhaled breathing of human beings contains a much higher concentration of carbon dioxide than the normal outdoor air. As a matter of consequence when persons are gathered in a room this leads to a noticeable increase of its concentration as it was recognized by previous authors [30]. Considering the situation depicted in figure SM2-1, a conservation equation for CO_2_ can be written in the same way than for particles or quanta:

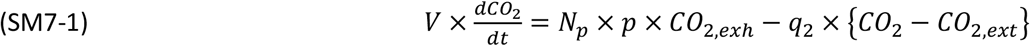

with the same notation meaning than in SM2 for *V, N*_*p*_, *p* and *q*_2_. *CO*_2_ is the current concentration of CO_2_ which can be expressed in ppm (part per million) since air density is assumed constant. *CO*_2,*exh*_ and *CO*_2,*ext*_ are respectively CO_2_ concentration in the air exhaled by a human (close to 40 000 ppm) and outdoor fresh air (around 420 ppm).

The last term of the equation comes from the fact that the fresh outdoor air contains CO_2_.

It follows that the carbon dioxide concentration in the room, equal to *CO*_2_(0) at *t* = 0, will evolve following the equation:

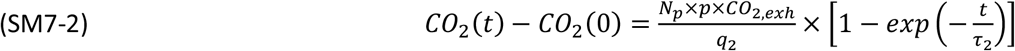

with

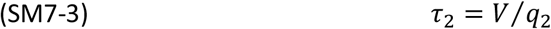

Note that most often a “clean” room with a null virus concentration *n*_*q*_(0) = 0 corresponds to *CO*_2_(0) = *CO*_2,*ext*_, excepted in un-stationary conditions, for example if ventilation is off during the night and considering a virus lifetime, see end of this SM.

When the quantum (virus) lifetime is very large, *τ*_1_ defined by equation SM2-4 reduces to *τ*_2_. Then, at any time *t*, it is straightforward to deduce from equations SM2-3, SM2-5 and SM7-2 that:

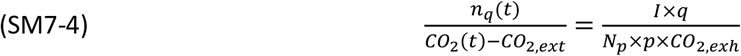

which assumes that at *t* = 0, *CO*_2_(0) = *CO*_2,*ext*_ and *n*_*q*_(0) = 0.

Note that it can be shown that the same equation holds for the poorly ventilated case developed in the main paper.

Then using the fact that the dose is:

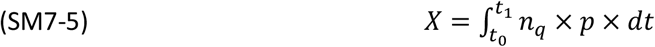

It follows, for a time of exposure Δ*t* = *t*_1_-*t*_0_, that:

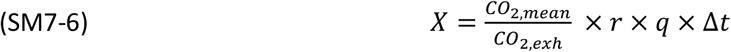

with:

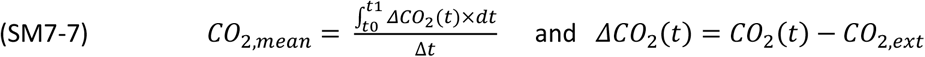

This equation was first established by Rudnick and Milton [30] in a different way and is valid even for unstationary conditions as long as the virus lifetime *τ*_*i*_ ≫ *V*/*q*_2_.

If the above conditions for *τ*_*i*_ is not fulfilled it is necessary to write a new equation for the dose as a function of time. Still assuming that at *t* = 0, *CO*_2_(0) = *CO*_2,*ext*_ and *n*_*q*_(0) = 0, Equation SM7-4 is changed as:

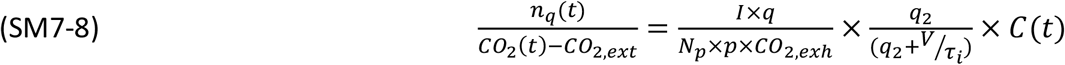

with:

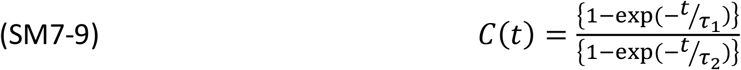

which at stationary state reduces to:

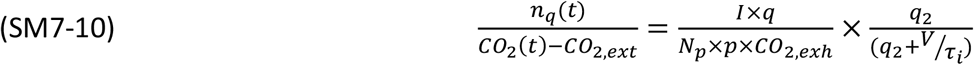

and for the dose at stationary state:

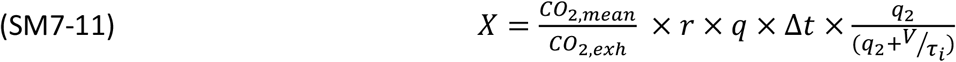

Of course, for transient evolution the dose can be calculated using equation SM7-5 with SM7-8. Now other transient evolutions could be considered with different initial conditions than the choice made above. For example if the ventilation is off overnight the virus lifetime could be such that in the morning (*t* = 0), the conditions *n*_*q*_(0) = 0 holds but with *CO*_2_(0) > *CO*_2,*ext*_. In this case *CO*_2,*ext*_ should be replaced by *CO*_2_(0) in equations SM7-8 and SM-7-10. These considerations show the importance of the virus lifetime, which is strongly dependent of the room conditions, especially the temperature [31]. Nevertheless, it remains that CO_2_ is most often an excellent proxy of the risk, excepted when an air sterilizer at high volume flow rate *q*_3_ ≥ *q*_2_ is used.

